# Unraveling the Link between CNVs, General Cognition, and Individual Neuroimaging Deviation Scores from a Reference Cohort

**DOI:** 10.1101/2023.11.29.23298954

**Authors:** Charlotte Fraza, Ida E. Sønderby, Rune Boen, Yingjie Shi, Christian F. Beckmann, Andre F. Marquand

## Abstract

Copy number variations (CNVs) are genetic variants that can have a substantial influence on neurodevelopment, neuropsychiatric traits, and morphometric brain changes, yet their impact at the individual level remains unknown. Common case-control approaches for analyzing CNVs suffer from limitations: they are unable to inform on individual variation between carriers and preclude the study of rarer variants, due to their limited sample size. This cross-sectional study aims to map individualized brain deviation scores in individuals with pathogenic CNVs. We used normative modeling to map neuroimaging features from several large neuroimaging datasets and applied these models to understand the neurobiological profile of CNV carriers in the UK Biobank. We highlight the 1q21.1 distal deletion and duplication, as an example of our individual-level normative modeling-CNV approach. Next, we counted the number of extreme deviations for each participant from the mean and centiles of variation from population reference norms, giving us a combined risk score per participant per imaging modality. We show a high degree of heterogeneity between pathogenic CNV carriers in their implicated brain regions. For example, the cerebellum, brainstem, and pallidum show large negative deviations for specific 1q21.1 duplication carriers. For certain 1q21.1 deletion CNV carriers the caudate and accumbens show notable positive deviations. Finally, we show that negative deviations from these models are correlated to cognitive function. This study marks a starting point in understanding the impact of pathogenic CNVs on brain phenotypes, underscoring the intricacies of these genetic variations at the individual level and providing a means to study the effects of rare CNVs in carrier individuals.

## 1. Introduction

Copy number variations (CNVs) are genetic variants that can have a large influence on neurodevelopment, neuropsychiatric traits, and morphometric brain changes (1,2). Some CNVs emerge as genetic risk factors for a variety of neurodevelopmental and other psychiatric disorders (3–7). Specifically, previous studies have shown that certain rare recurrent CNVs increase the risk for schizophrenia (5,8–12), attention deficit hyperactivity disorder (ADHD) (13), autism spectrum disorder (ASD) (14,15), and links between a decreased intelligence quotient (IQ) score and pathogenic CNVs have been established (16). Despite their known importance, the effects of the majority of individual CNVs on the brain and behavior remain largely unknown. CNVs are typically studied at the group level, which may mask considerable inter-individual variation in their brain phenotypic presentation. Furthermore, the group-based approach is only feasible for the more common CNVs, limiting our ability to map the effects of rarer CNVs with high penetrance or large effect sizes. Understanding the effects of specific CNVs on both brain structure and cognition at the individual level is crucial for mapping their impact on mental disorders.

CNVs often have pleiotropic effects, influencing multiple downstream processes simultaneously (17–19). The diversity in genetic CNV-mediated effects is heightened by interactions with both the rest of the genome and environmental factors (20). This makes each CNV’s impact unique to the individual, complicating the task of understanding their contributions to overall mental health. Conventional imaging and behavioral CNV studies often adopt a cases vs. controls framework, which has led to tremendous insights. However, in order to grasp the full spectrum of CNV effects, which manifest in a quite heterogeneous manner, we need to move beyond group-level distinctions (21).

Understanding the individual-level impact of CNVs on the brain and behavior has posed a challenge, yet the emergence of large-scale normative models may offer a solution. By employing this approach, z-scores can be calculated for each individual across various neuroimaging modalities, quantifying individual variations against the mean and centiles of population reference norms. Normative models thereby shift focus from group-level to individual-level inferences (22–25) and allow us to quantify atypical developmental trajectories (26,27). Normative modeling has proven its efficacy in correlating individual behavioral phenotypes with deviations from reference cohorts, spanning disorders like schizophrenia (26,28), neurodivergence for ASD and ADHD (29,30), and mapping disease progression in Alzheimer’s disease (31). Using normative models, we can map brain deviation scores for individuals with a specific pathogenic CNV. Afterward, we can use these pathogenic CNV-brain phenotypic deviation scores and correlate them with behavioral traits, uncovering hidden facets that group analyses could potentially miss. Importantly, owing to its focus on individual differences, this approach is not limited to common CNVs; it extends to rarer variants, opening doors to tailored research for smaller populations of rare genetic variants.

The normative modeling approach is specifically designed to give insight into individual brain deviation scores and their relationship to mental health. Mental health disorders such as major depression and schizophrenia lie on a gradient of severity and can be seen as the extreme values on continuous dimensions (32). This idea aligns with normative modeling, as it places individuals with large brain deviation scores at the edges of the normative spectrum (33). In this study, our primary objective is to dive into the individualized impacts of CNVs on brain structure and behavior. By using a normative modeling approach, we take a step towards making personalized risk profiles that allow us to do cross-individual comparisons. These profiles prove insightful for individuals with similar genetic mutations that manifest in comparable behavioral effects and share diverse neuroimaging fingerprints. We hypothesize that: (i) individuals with a CNV related to cognitive deficits or neurodevelopmental disorders, will have larger deviation scores compared to a reference model across many brain areas and (ii) that the patterns of deviation across brain regions will be highly variable across these CNV carriers.

## Methods and materials

Ethical approval for all the data used in this study was attained previously by the studies contributing their datasets. Details of all the data can be found in the publicly available repositories’ main publications: Cam-CAN (34), HCP (35), Oasis (36), PNC (37), and UK Biobank (38).

### From pathogenic CNVs to brain structures: a normative modeling approach

An overview of our analytic workflow is presented in Fig. 1 and a flowchart of the two data samples used in the subsequent analysis is visualized in Fig. 2. All the details of the different analyses described here can be found in the supplement. Additionally, the models used for all analyses are accessible at https://github.com/amarquand/PCNtoolkit. In brief, to discern whether certain neuroimaging modalities exhibited more atypicalities than others, we first employed normative modeling to analyze the pre-estimated Image Derived Phenotypes (IDPs) from the UK Biobank study (38), covering diverse functional, structural, and diffusion tensor imaging measures. In total, we used data from 44,456 participants from the UK biobank and 2084 IDPs, which were processed using FUNPACK (39), developed by the Wellcome Centre for Integrative Neuroimaging and used for automatic normalization, cleaning, and parsing. From this initial study, we found that the IDPs derived from the structural measures contained the largest variation between deviation scores among participants, see also the result section.

**Fig. 1.**
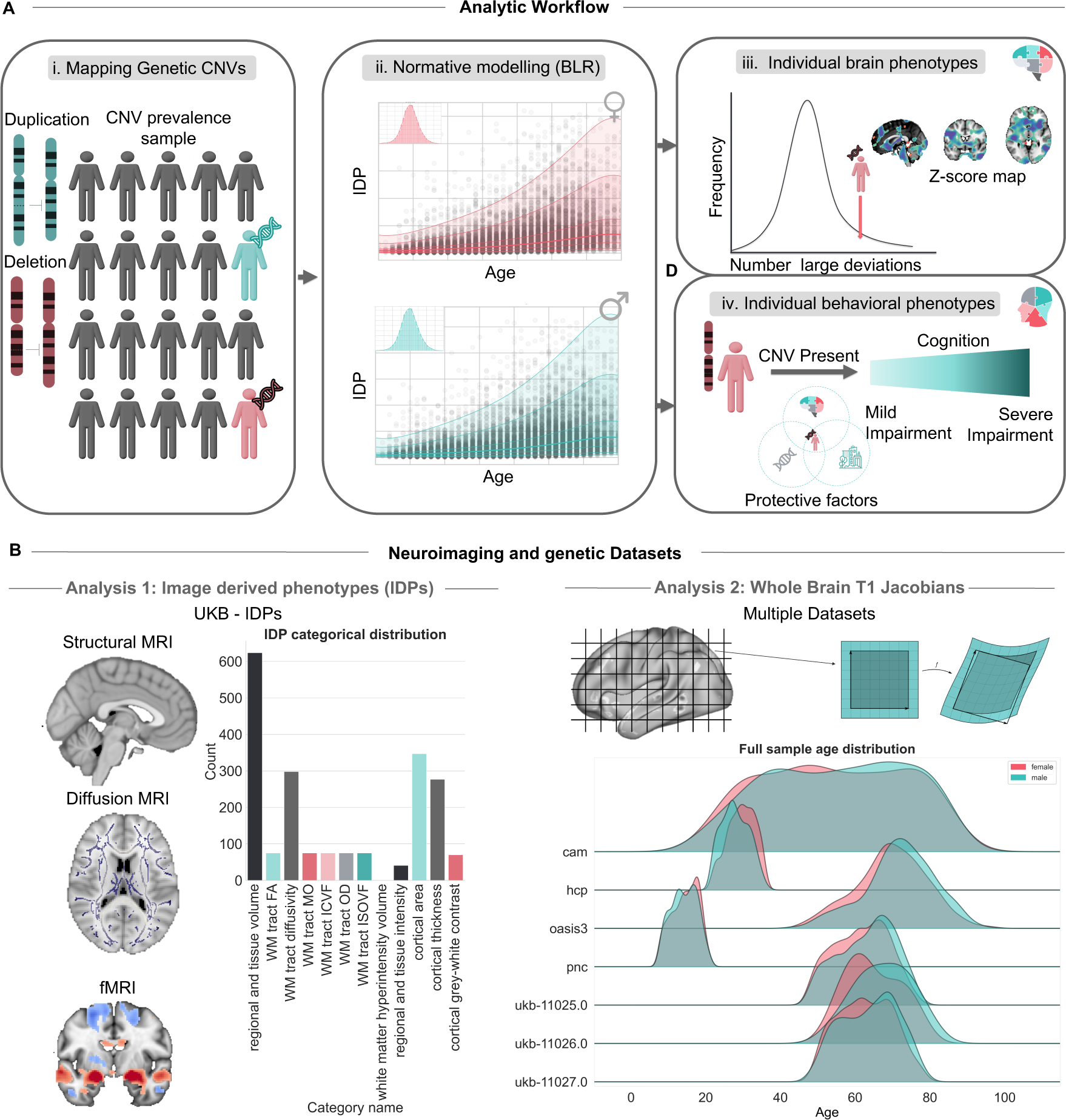
| Overview of study design and data resources. **A**. Schematic overview of the study workflow and hypothesis. First, we quantify the number of participants with pathogenic CNVs, previously linked to neurodevelopmental and psychiatric disorders. Then we create normative models for the IDPs and afterwards the voxel-wise Jacobians. We calculate the number of large deviation scores (|Z|>2). We plot the number of large deviation scores for individuals with pathogenic CNVs compared to the rest of the population. Finally, we correlate the extreme brain deviation scores of the Jacobian measures with a general cognitive ability score. CNV, Copy Number Variant; IDP, image derived phenotype; BLR, Bayesian Linear Regression. **B.** Overview of the neuroimaging datasets used in this study. i. Distribution of the IDPs present in the UK biobank dataset, derived from functional, structural, and diffusion tensor imaging. ii. Distributions of the data from seven sites used in the Jacobian normative model, split by sex. In total, for the IDP study, we use 43,893 participants, and for the Jacobian-voxel-based study, we use 19,620 visually quality-controlled participants.

**Fig. 2.**
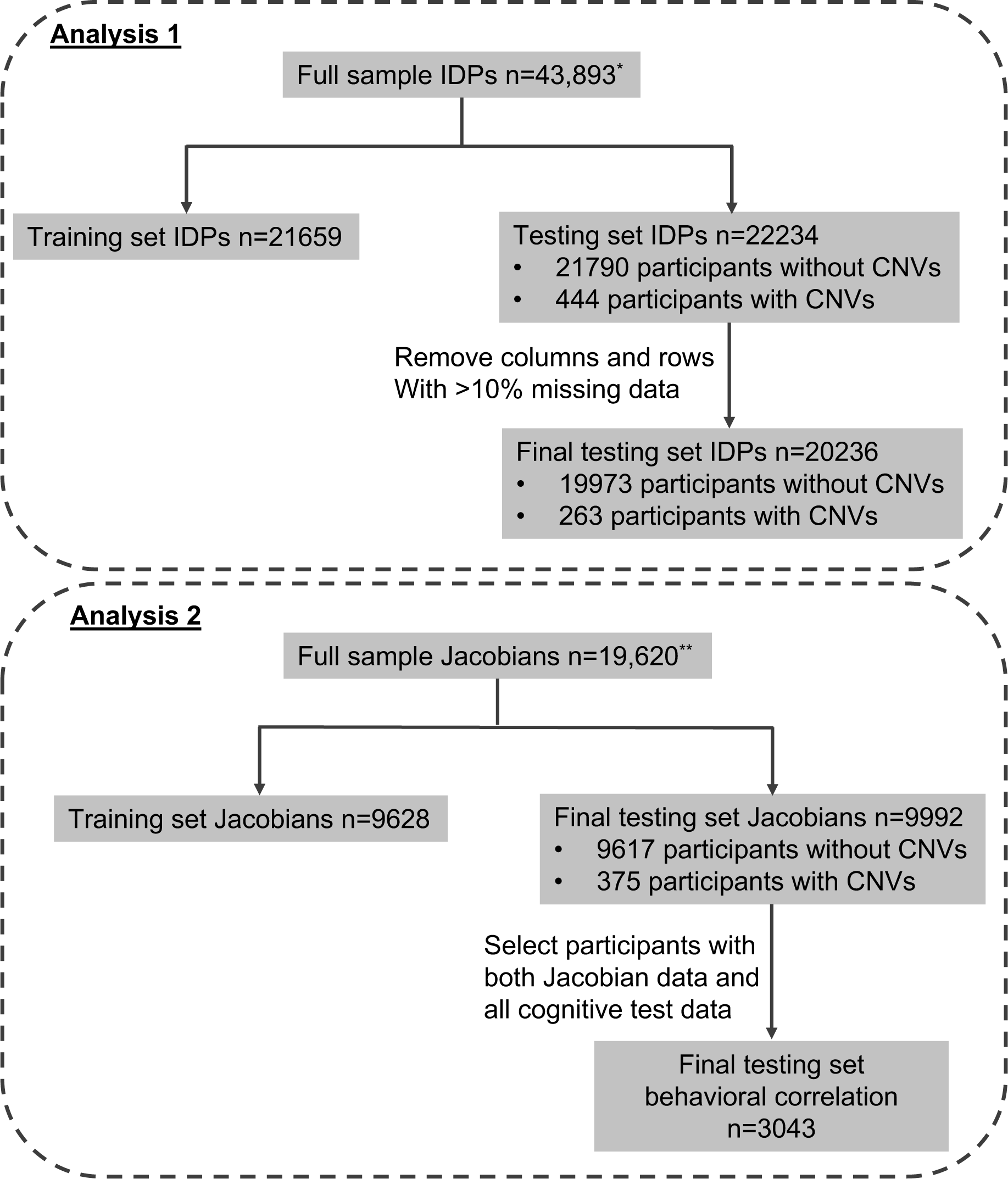
| Flowchart number of individuals used at each stage of the study. We report the number of participants for analysis one, focusing on Image Derived Phenotypes (IDPs), and analysis two, concerning whole brain T1 Jacobians. *Quality control for the IDPs was performed previously by the UK Biobank (38). **Visual quality control for the reference Jacobian datasets has been performed previously and detailed in the following study (24).

To take this result forward we wanted to explore a whole-brain voxel-based model based on structural T1 data and to map in a spatially precise manner the individual-level effects of pathogenic CNVs within the brain. More specifically, we fit a voxel-based morphometric variation model using Jacobian determinant images derived from the non-linear image registration to the MNI152 space. Jacobians, in their essence, provide a spatially precise measure of voxel-based morphometric differences, capturing the extent of volumetric adjustments—either expansion or contraction—needed to align each sample with the registration template for each voxel. These determinants are more informative compared to other derived measures (40) and describe aggregate differences, avoiding the partly arbitrary distinction between grey and white matter. Moreover, it is well-established that specific CNVs influence intracranial volume (ICV) (41,42). Consequently, we anticipated that this influence would manifest as either an increase or decrease in the necessary volumetric adjustments for individuals with these CNVs, which would be reflected in their Jacobians. Typically, a normative model can be constructed based solely on the age range of interest. In our case, we would then be focusing on the 40-80 age bracket where CNV carriers are located. However, it is advisable to develop a normative model covering the entire age spectrum. This ensures that we make an accurate estimation even at the edges of the age range of interest, by aligning the estimated centiles with a wider age range and thereby reducing uncertainty in the centile estimations. To create the voxel-wise Jacobian normative model that spans the entire age range, we pooled a large dataset from five publicly available repositories: Cam-CAN (34), HCP (35), Oasis (36), PNC (37), and UK Biobank (38), leveraging Jacobian determinant images from non-linear image registration, specifically via the anatomical processing tools found in FSL. The details of the quality control, preprocessing steps, and data resources can be found in the supplement. In total, we used visually quality-controlled T1 Jacobian data from 19,620 participants from seven sites. To indicate how representative the samples are, in Tables 1 and 2, we present an overview of the demographics of the samples employed in this study. The ethnicity data presented in the studies originates from self-reported data from the publicly available datasets from which the samples were derived. We decided to use the same ethnicity categories as the UK Biobank, according to: https://biobank.ctsu.ox.ac.uk/crystal/field.cgi?id=21000, which included the categories W = White, B = Black, A = Asian, M = Mixed and O = Other. These categories are quite general and therefore heterogenous in the populations they represent, for example, the label Asian can indicate both South and East Asian backgrounds.

**Table 1.** | Demographics for analysis 1 (Image Derived Phenotypes) The ethnicity data is collected as self-reported data by the UK biobank with the labels M=Mixed, A=Asian, B=Black, C=Chinese, W=White, and O=Other.

| Sample | N | Sex<br>(F%/M%) | Mean age<br>(SD) | Age range | Ethnicity (%) |
| --- | --- | --- | --- | --- | --- |
| **UKB-IDP** | 43,893 | 52.4/47.6 | 60.97 (8.72) | 44-82 | 96.75 W/ 0.44 M / 1.05 A / 0.65 B/ 0.30 C/ 0.53 O |
| **UKB-IDP-CNV** | 263 | 54.4/45.6 | 62.97 (8.70) | 46-80 | 96.96 W/ 1.14 M/ 1.14 A/0.00 B/0.38 C/ 0.00 O |

**Table 2.**
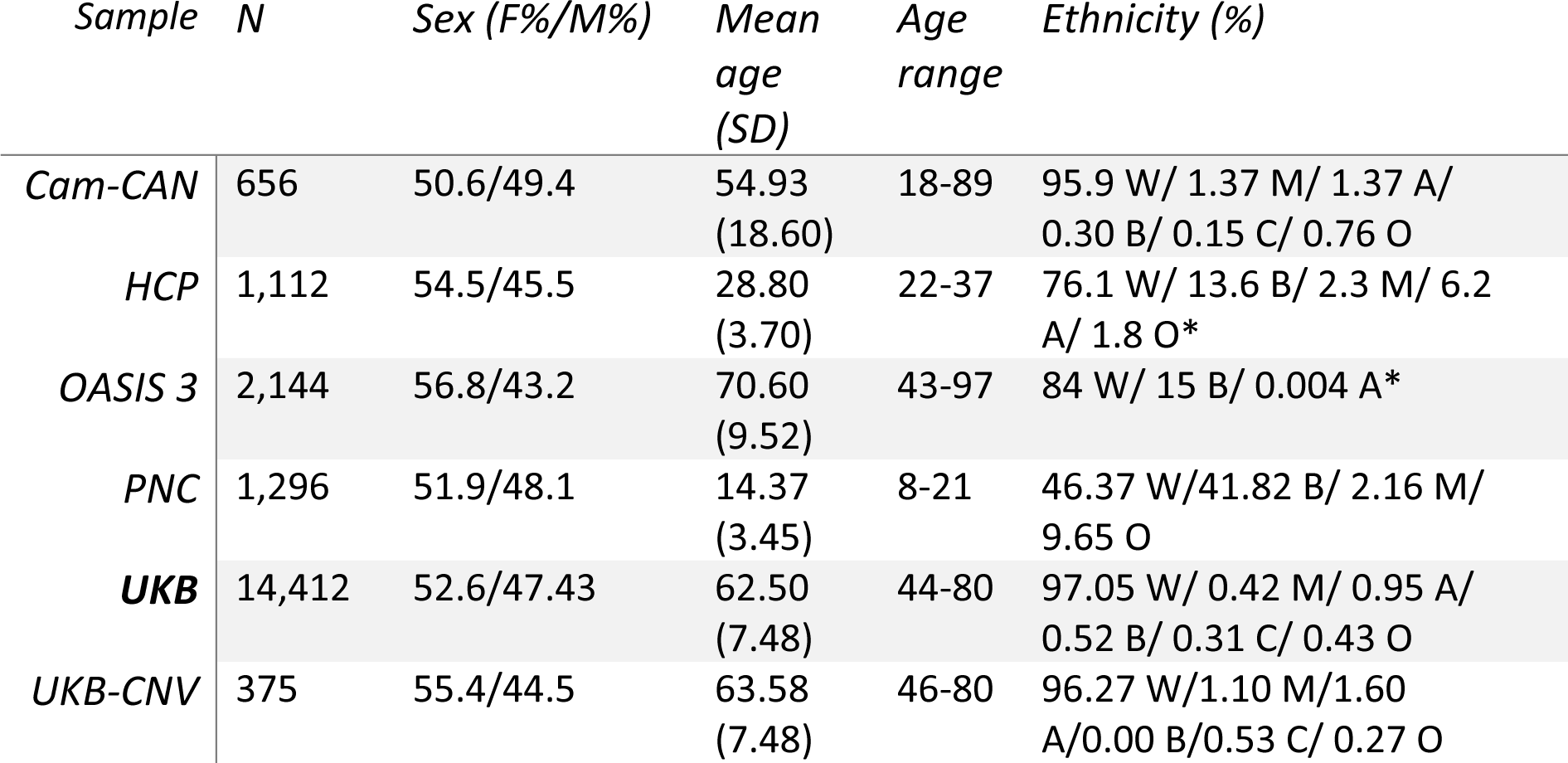
| Demographics for analysis 2 (whole brain T1 Jacobians) The ethnicity data is summarized with the labels M=Mixed, A=Asian, B=Black, C=Chinese, W=White, and O=Other. *Ethnicity estimation was derived from previous work for the HCP sample in (60) and for the OASIS 3 sample in (61).

We employed the Bayesian Linear Regression (BLR) function from the PCNtoolkit to create the normative models for each voxel, the mathematical details are described in the supplement. To ensure accurate modeling of non-linear and non-Gaussian effects, we employed a B-spline basis function and likelihood warping, described in (43). Every normative model constructed factored in the covariates age and sex. To reduce site effects, we have added it as a fixed effect in the model. In general, incorporating site as a confounding variable during the fitting procedure is the most effective approach for handling site effects (44). Harmonization procedures can also be considered, if the site data is unavailable or for adapting larger models afterward. However, they come with extensive drawbacks, including the potential removal of meaningful variance correlated with site effects (45,46).

We focussed on 92 CNVs proposed to be pathogenic (henceforth ‘pathogenic’), and their reciprocal CNVs (47–49). These can be requested from https://biobank.ndph.ox.ac.uk/ukb/app.cgi?id=14421. We highlighted the 1q21.1 distal deletion and duplication, as an example of our normative modeling-CNV approach. This CNV has shown moderate to strong effects on cognition (4,48), a dose-response per copy number for head circumference (50), with microcephaly in deletion carriers and macrocephaly in duplication carriers, and has been associated with global cortical surface structure changes (41). Furthermore, individuals with a 1q21.1 deletion and duplication show an increased risk for several neurodevelopmental disorders (3,4,50–52). The choice of focusing on one CNV gives us the ability to give a full overview and visualization of the implications specifically associated with this CNV. Nonetheless, we have also included results for three other CNVs—15q11.2 deletion and duplication, 16p11.2 deletion and duplication, and 16p13.11 deletion and duplication—in the supplementary materials for interested readers. These CNVs were selected based on prior research indicating their impact on neurodevelopment and cognition (21,53,54). While our study is primarily designed to unravel individualized pathogenic CNV effects, we also explored the potential for aggregating subjects with similar CNVs to uncover converging brain structural alterations across pathogenic CNVs. This will allow us to also make comparisons with traditional case-control studies.

Finally, we linked the found Jacobian brain deviation scores to cognition by testing a Spearman correlation between the number of extreme deviation scores (|Z|>2) per participant and a general measure of cognitive ability, derived from seven cognition tests available in the UK biobank (https://biobank.ndph.ox.ac.uk/ukb/label.cgi?id=100026) (55). Our choice to focus on cognition scores as the primary behavioral variable stemmed from prior research indicating that the selected CNVs may detrimentally affect cognition (56,57). Additionally, for those interested, we included a supplementary behavioral correlation analysis linking all IDP deviation scores and Jacobian deviation scores to several other behavioral variables sourced from the UK Biobank. Furthermore, to test the correspondence between the IDP and Jacobian deviation scores with the polygenic scores (PGS) for major neuropsychiatric disorders, we included a correlation analysis with the PGS for seven disorders: ADHD, ASD, major depressive disorder (MDD), anxiety disorders (ANX), schizophrenia (SCZ), bipolar disorder (BIP), and cannabis use disorder (CUD). The PGS scores were calculated with the PRS-CS-auto method (58). Details of the PGS derivation and correlation analysis can be found in the supplement and the related paper (59).

Fig. 1A visually outlines our study workflow: i. We first mapped participants with pathogenic CNVs in the UK Biobank and placed them in our test set of the normative models. ii. We created normative models for each individual IDP or voxel, taking into account covariates age, site, and sex. iii. We counted the number of extreme deviations for each participant (|Z|>2) and mapped where participants with a pathogenic CNV lay on this distribution, giving us a combined risk score per participant per imaging modality. iv. We correlated the total number of extreme Jacobian z-scores with a measure of general cognitive ability. In Fig. 1B, we show an overview of the datasets used, encompassing IDPs from the UK biobank and Jacobian data from seven sites, details can be found in the supplement. In total, we used 44,456 participants in the IDP study, and in the Jacobian-voxel-based study 19,620 participants that passed visual quality control. There were 375 individuals with pathogenic CNVs in the quality-controlled Jacobian neuroimaging dataset. In Fig. 3A and 4A, the final pathogenic CNV sample used for the Jacobian normative model study is shown.

**Fig. 3.**
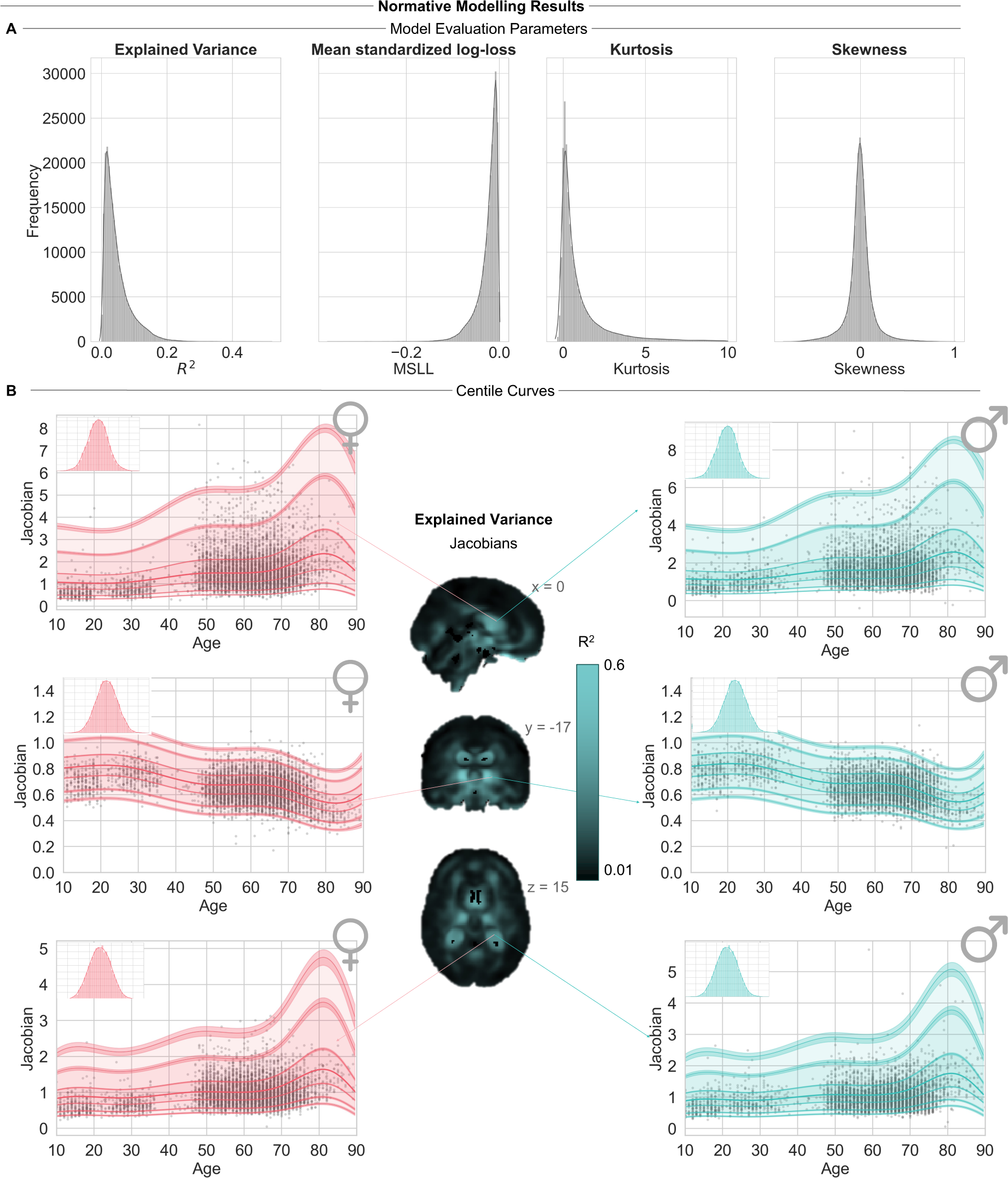
| Overview of normative modeling results. **A**. Performance metrics for the test set. Both skew and kurtosis serve as indicators of the model’s accuracy in estimating shape via warped Bayesian Linear Regression. **B**. Depiction of varied normative trajectories across distinct voxels, showing in the corner the histogram of z-scores, with the accompanying whole brain explained variance (*R*^2^) map, split based on sex.

**Fig. 4.**
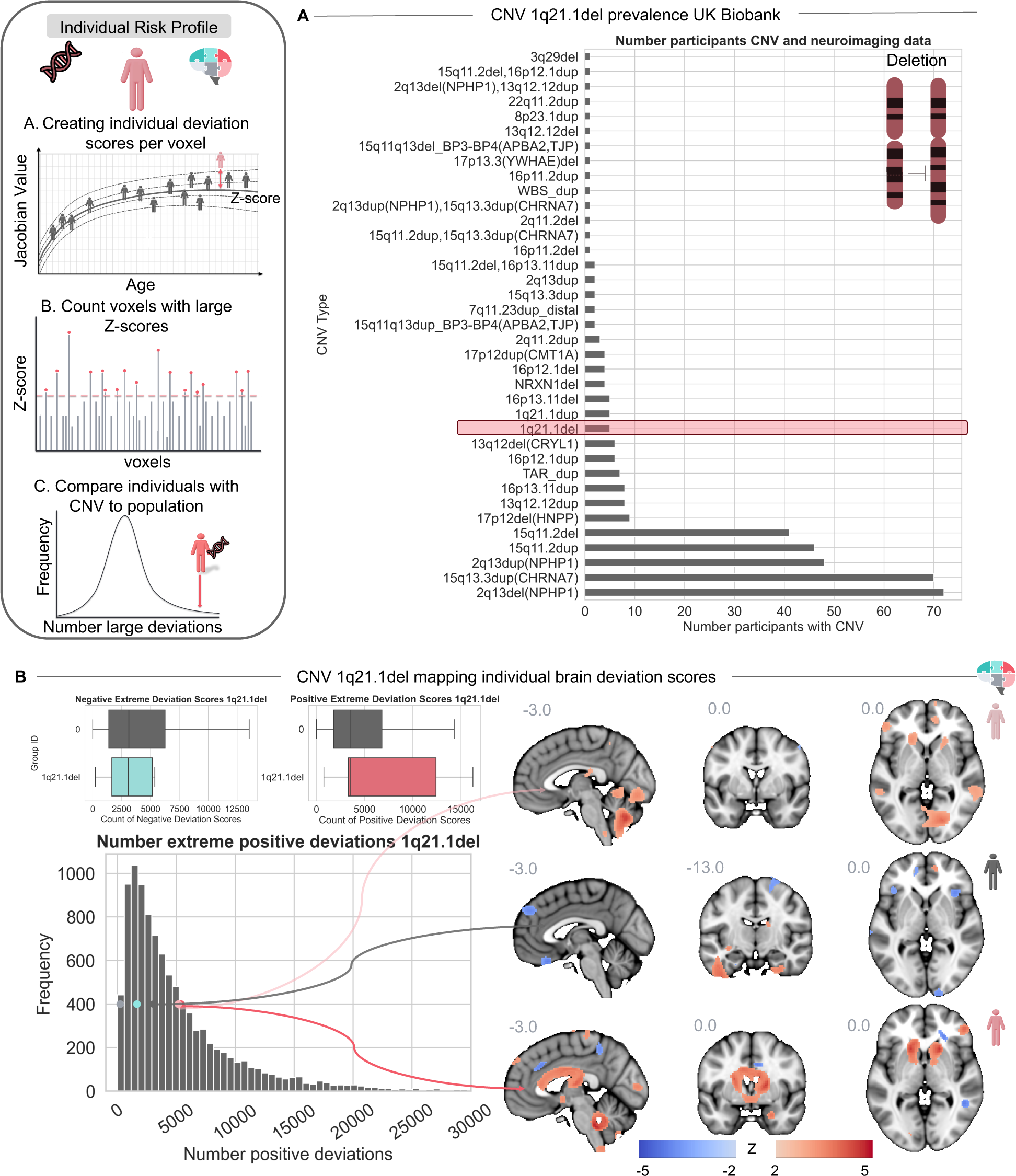
| Individual risk profiles 1q21.1 deletion. **A.** The prevalence of pathogenic CNV carriers including 1q21.1del in the UK Biobank neuroimaging dataset used in Jacobian analysis. **B.** Counts of extreme positive and negative deviation scores (|Z|>2) among participants with a 1q21.1 deletion in contrast to participants without a pathogenic CNV. Left: Dots show each individual 1q21.1 deletion carrier’s position in the distribution. Right: Displaying the profile of three selected 1q21.1 distal deletion CNV carriers.

## Results

Initially, we fitted a normative model to the IDPs derived from the structural, functional, and diffusion measures from the UK biobank. The IDPs give global summary measures for each modality and have been previously publicly released (38). Specifically, we were interested in the differences between participants who had pathogenic CNVs in comparison to those without. Detailed results from this IDP normative model can be explored in the supplementary Fig. 1, 2, and 3. The resulting models explained as much as 45% of the variance, according to the maximum of : 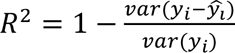, with *y_i_* the true values and *ŷ_i_* the predicted values, using the covariates age, sex, and site. By considering these covariates, any residual variation in the model is largely uncorrelated with them and is likely attributable to other variables. A notable observation was that structural measures appeared most informative when we explored individual deviation scores. In supplementary Fig. 2 and 3, we can see that the structural measures gave a larger spread of the number of extreme deviations compared to other measures, which indicates a larger variation in individual differences.

Recognizing the importance of structural measures, we mapped a voxel-based morphometric variation model to characterize these differences at a finer scale, see Fig. 2. The resulting models accounted for as much as 52% of the variation in morphometric changes, using the covariates age, sex, and site. To validate the models we used several model fit criteria. Among these were the kurtosis and skewness of the resulting z-score distribution, which measures the effectiveness of the warping function in capturing the non-linearity and non-Gaussianity of the data (43). In general, all voxels show relatively low skew (i.e. |skew|< 1) and acceptable excess kurtosis (<5).

### Individual risk profiles - combining pathogenic CNVs and brain imaging deviation scores

Next, we aimed to characterize the individual variations in brain deviation scores associated with specific pathogenic CNVs. To accomplish this, we counted the number of extreme deviations (|Z|>2) for each individual. An overview of the average counts of negative and positive deviation scores across all pathogenic CNVs is presented in supplementary Fig. 4.

Afterward, we constructed individualized risk profiles for 1q21.1 distal deletion (1q21.1del) and duplication (1q21.1dup). We chose this CNV to highlight our method, as it is relatively well-characterized with regard to brain structural differences (21), has shown impairments in cognition, and has been associated with neurodevelopmental disorders (50). Fig. 3 and 4 depict the number of positive and negative deviation scores for individuals with a deletion or duplication, respectively, in comparison to participants without pathogenic CNVs. Additionally, these figures showcase deviation score brain maps for individuals with a pathogenic CNV, highlighting regions that have pronounced positive or negative deviations when compared to a reference cohort, demonstrating a high degree of variability of volumetric alterations across carriers. The maps show areas with marked negative or positive Jacobian signals or localized volumetric alterations. The interpretation of positive and negative deviations can vary depending on the mean of the normative model. For example, when the deviation is positive compared to the mean, it indicates more expansion and a relatively small volume in that voxel compared to the norm. We can see from panel B that positive deviations, i.e. more volume expansions than predicted, were more present for participants with a 1q21.1del, and negative deviations, i.e. more volume contractions than predicted by the model, were more prevalent for participants with a 1q21.1dup. For the detailed brain maps of all the participants with a 1q21.1 distal deletion or duplication, see supplementary Fig. 5 and 6. The most implicated brain areas, calculated using the mean deviation score per ROI, are also summarized in word clouds.

Then, we conducted a joint analysis for the 1q21.1del and 1q21.1dup groups, respectively, to examine the common distribution of deviation scores across all subjects. We expect that there is convergence in deviation scores amongst the individual subjects in certain brain regions (54), especially those that can be found with traditional cases vs. control paradigms, but also we expect divergence amongst the subjects in deviation scores in other brain regions, as we know that not every participant with a pathogenic CNV is implicated in their behavioral phenotype or cognition. For example, a participant can have a pathogenic CNV and still have a standard cognitive performance. The analysis revealed prominent deviations in specific brain regions, see Fig 5. Notably, for 1q21.1dup, substantial negative deviations can be seen in the occipital cortex, while for 1q21.1del, pronounced positive deviations were observed in the cerebellum and thalamus. In supplementary figures 9-14, individual risk profiles for the pathogenic CNVs 15q11.2 deletion and duplication, 16p11.2 deletion and duplication, and 16p13.11 deletion and duplication are presented, showcasing the versatility of our approach, which can be applied to diverse rare pathogenic CNVs.

**Fig. 5.**
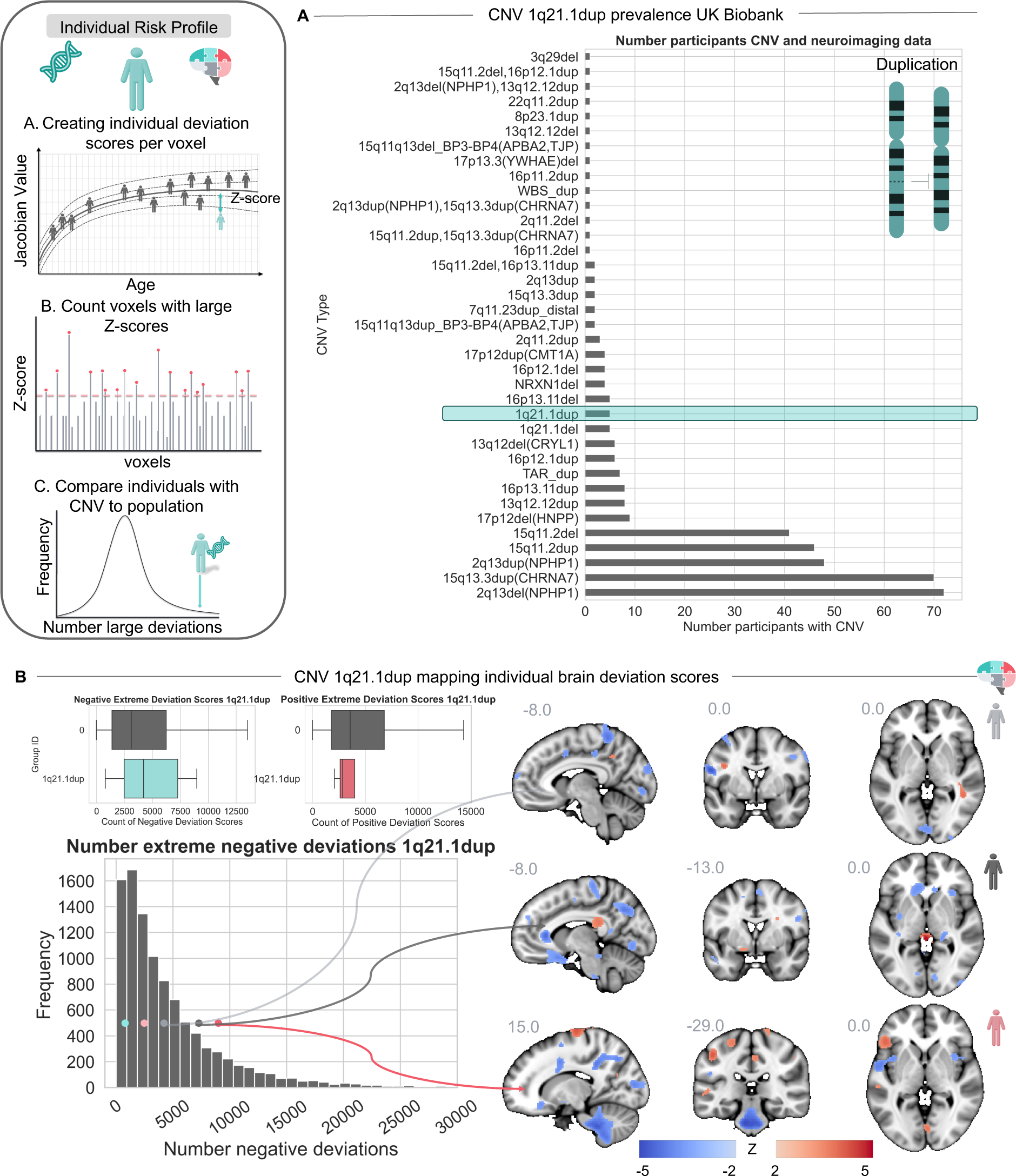
| Individual risk profiles 1q21.1 duplication. **A**. The prevalence of pathogenic CNV carriers including 1q21.1dup in the UK Biobank neuroimaging dataset used in Jacobian analysis. **B.** The counts of extreme positive and negative deviation scores (|Z|>2) among participants with a 1q21.1 duplication in contrast to participants without a pathogenic CNV. Left: Dots show each individual 1q21.1 duplication carrier’s position in the distribution. Right: Displaying the profile of three selected 1q21.1 distal duplication CNV carriers.

### Relationship brain deviation scores and cognitive deficits

Fig. 6 outlines our hypothesis concerning the impact of pathogenic CNVs on cognition, proposing that certain CNVs might contribute to cognitive impairment in the absence of protective factors. We plotted the fluid intelligence scores among participants with pathogenic CNVs and participants without pathogenic CNVs in Fig. 6A. To analyze the relationship between large deviation scores and cognition, we generated a general cognitive ability score for each participant by calculating the first principal component from the various cognitive tests within the UK Biobank dataset, see the supplementary figure 7 for an overview of the tests used. We examined the Spearman correlation between the total count of extreme positive deviation scores (Z>2) and the general cognitive ability score (r = 0.03, p = 0.09) and the extreme negative deviation scores (Z <-2) and the general cognitive ability score (r = −0.04, p = 0.04), see Fig. 6. This correlation indicated that a higher number of extreme negative deviations, indicating more volume contractions compared to the mean of the population, was significantly associated with a lower general cognitive ability score. In supplementary figures 18-22, we included the outcomes of Spearman correlation analyses between the total count of extreme positive and negative deviation scores and various other behavioral variables available in the UK Biobank, for the interested reader.

**Fig. 6.**
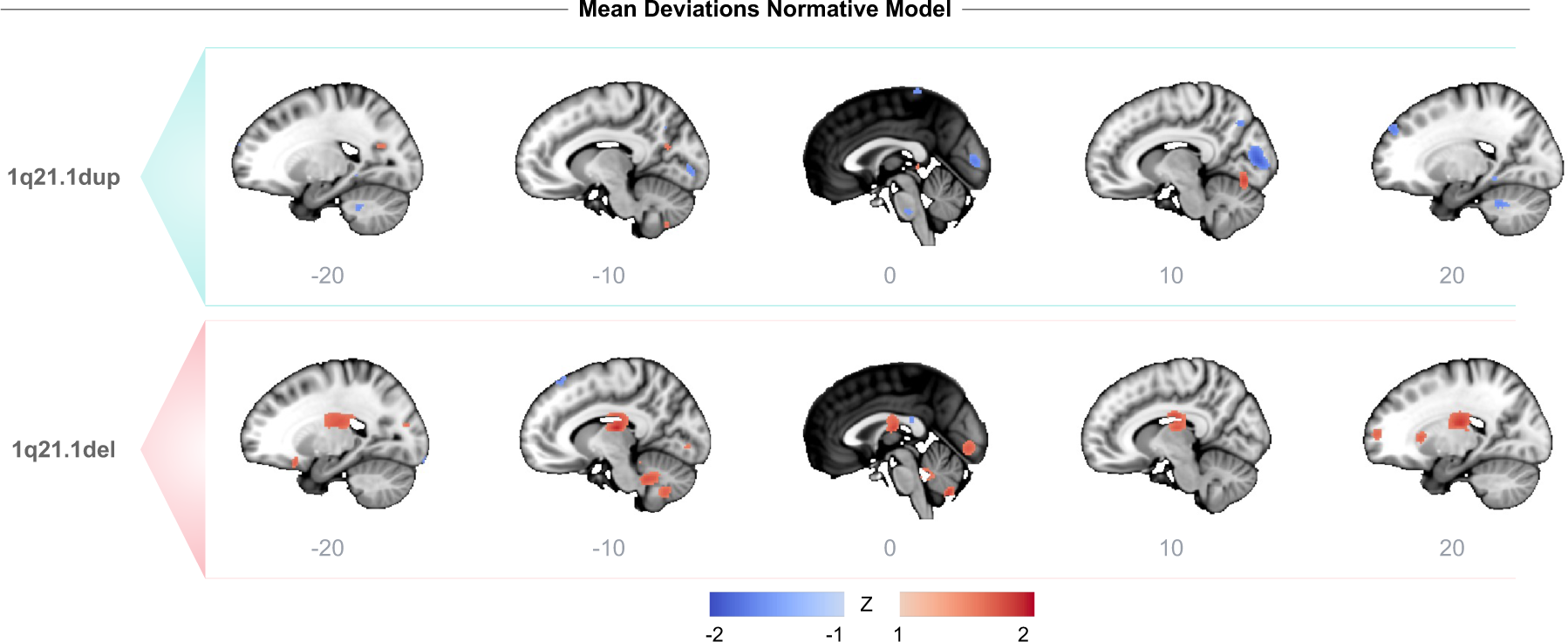
| Convergence of positive or negative deviation scores. The mean brain deviation score maps for participants with the 1q21.1 distal deletion or duplication CNV. On the x-axis, we show different sagittal slices with steps of 10.

**Fig. 7.**
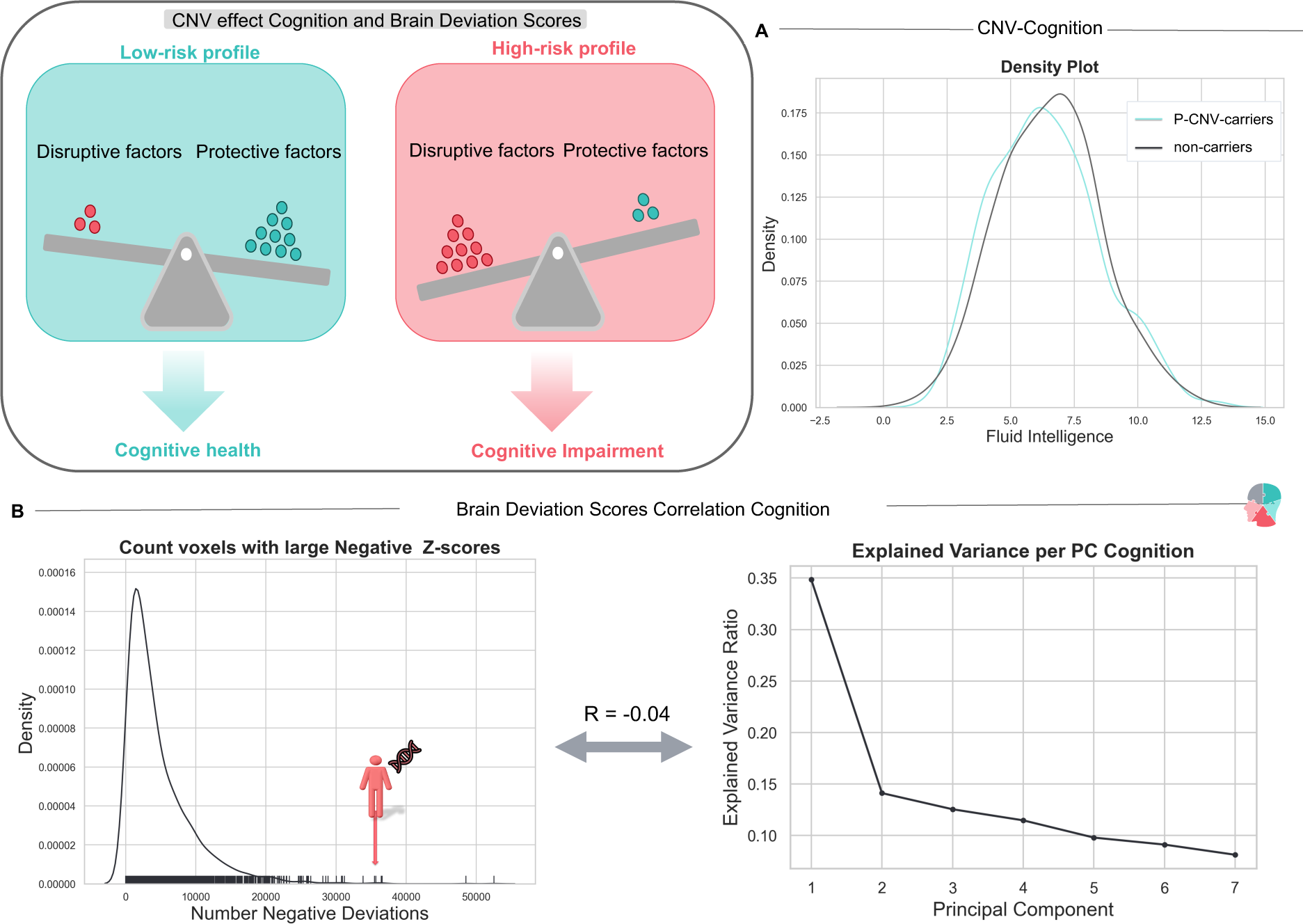
| Pathogenic CNVs impact on cognitive function. Illustration of how protective and disruptive factors, which may include (pathogenic) CNVs, might lead to cognitive impairment. **A.** Variations in fluid intelligence scores among participants with pathogenic CNVs (P-CNVs) and without pathogenic CNVs (non-carriers). **B**. Depicting the found Spearman correlation between the number of large negative deviation scores (Z<-2) and the first principal component of cognition, which is usually called general cognitive ability.

## Discussion

### Individualized risk profiles through normative models -

The aim of this study was to characterize individualized risk profiles for participants with pathogenic CNVs related to neurodevelopmental and psychiatric disorders. In this endeavor, we are the first to concurrently map individualized brain deviation scores in individuals carrying pathogenic CNVs. Our study took advantage of the UK Biobank, enabling us to analyze multiple pathogenic CNVs concurrently. We employed a normative modeling approach to generate individual difference maps for both IDPs and detailed Jacobian measures indicating volumetric changes at the voxel level and the level of the individual. Initially, we established normative reference models from IDPs to identify the neuroimaging modality most influenced by the pathogenic CNVs. Our findings highlighted that structural measures appeared more informative. Following this, we utilized a whole-brain Jacobian model to map deviation scores with voxel-wise precision.

In the introduction, we posed two main hypotheses: (i) individuals with a CNV related to cognitive deficits or neurodevelopmental disorders will have larger deviation scores compared to a reference model in certain brain areas, and (ii) the patterns of deviation across brain regions will be highly variable across these pathogenic CNV carriers. To confirm these hypotheses, we analyzed the deviations from the reference model’s normative trajectories, by binning the deviation scores. We counted the number of extreme volume contractions (Z<-2) and expansions (Z>2), as compared to the mean and variance of the reference model. This methodology supplied us with an individualized map of extreme deviation scores. For each participant with a pathogenic CNV, this allowed us to pinpoint the brain regions showcasing the most pronounced deviations. Expanding on this, we counted the total positive and negative deviation counts and juxtaposed them against the broader population’s risk scores. As we expected, certain participants with a pathogenic CNV displayed an elevated number of positive or negative deviations compared to the general population, while others demonstrated deviation scores that aligned more with the ‘norm’.

Finally, we examined how the deviation scores are associated with general cognitive ability. We chose cognitive ability as our primary behavioral measurement because it has been consistently shown to be impaired within individuals with certain pathogenic CNVs (16,62) and thus allows us to establish a biologically meaningful connection between brain deviation scores and impaired behavior. Our analysis revealed a significant negative correlation between the count of negative deviations and general cognitive ability.

Using normative models to understand how pathogenic CNVs affect the brain has important benefits. It helps uncover hidden insights that group analyses might miss. Importantly, this approach focuses on individual differences, so it’s not limited to common CNVs but can be applied to rarer variations, making it possible to do research for smaller groups with uncommon genetic variants. For each person with a common or rare pathogenic CNV, we can identify the specific brain areas where they differ significantly from the norm. This allows us to see which brain regions are affected in each case, going beyond studies that need large sample sizes and can only look at more common CNVs.

### Individualized risk profiles for 1q21.1 distal CNV -

We made individualized risk profiles for both carriers of the 1q21.1 distal deletion and duplication from the deviation scores. Participants with this CNV exhibit a diverse range of impaired traits (56,57), which aligns with our proposal of an individualized approach. Notably, 1q21.1 CNV is associated with several, different neurodevelopmental disorders (2,4). For participants with a 1q21.1 deletion or duplication, we counted instances of extreme positive and negative deviation scores, contrasting these frequencies with the broader population. Subsequently, we generated brain maps alongside these deviation scores to highlight the regions of the brain where deviations were most prominent. We aggregated the deviation scores across participants with duplications and separately for those with deletions. This allowed us to identify brain regions where the effects converged across individuals and we can then subsequently compare it with previous literature that uses case-control setups.

For the Jacobian normative model, we quantify individual deviations from the mean volumetric change for a specific voxel. In this context, negative deviations refer to instances where certain brain regions show more volume contractions relative to the mean value of the Jacobian normative model. Put simply, these deviations might indicate that a specific brain region had a larger volume originally than what the normative model predicts for a typical voxel. The Jacobian image subsequently corrects for these differences. Positive deviations refer to cases where certain brain regions exhibit more volume expansions relative to the mean value of the Jacobian normative model. These deviations could represent an original lower brain volume in certain voxels compared to the predicted or typical voxel. Interestingly, individuals with a 1q21.1 duplication showed more negative deviations, thus these individuals on average have more volume contractions than expected by the model, reflecting larger intracranial volume and macrocephaly (41) in 1q21.1 distal duplication carriers. In contrast, 1q21.1 deletion carriers showed more positive deviations, which indicates that these participants have relatively more voxels with a lower brain volume compared to the mean of the population, which might reflect their smaller intracranial volume and microcephaly (41). In other words, we identified more positive deviation scores in deletion carriers and more negative deviation scores in duplication carriers, reflecting previous findings of dosage effects on the brain of the 1q21.1 distal carriers.

Previous literature has also revealed various effects associated with the 1q21.1 distal CNV on the brain, including positive dosage effects on ICV and total cortical surface area, particularly in the frontal and cingulate cortices, and negative dosage effects on caudate and hippocampal volumes (41). Another study found higher intraindividual variability in brain structure in 1q21.1 distal CNV carriers, with distinct regional effects on cortical surface area and thickness. Additionally, 1q21.1 distal deletion carriers exhibited reduced global cortical surface area, impacting primarily frontal and association cortices (21). Moreover, this CNV is linked to a high prevalence of micro- and macrocephaly in deletion and duplication carriers, respectively (56,57). From our results, we can see that the dosage effects of the 1q21.1 distal carriers remain the same. This means that a duplication of the copy number of the 1q21.1 region is associated with more negative deviations in the Jacobian, which indicates an increase in volume of certain brain structural features, and the 1q21.1 deletion is associated with more positive deviations in the Jacobian, indicating a relative decrease in volume of certain brain regions.

In general, our study shows different regions that are implicated for different participants, for example, the cerebellum, brainstem, and pallidum show large negative deviations for certain 1q21.1dup carriers. For certain 1q21.1del CNV carriers, the caudate and accumbens show significant positive deviations. A recent multivariate analysis of eight CNVs revealed that the cingulate gyrus, insula, supplementary motor cortex, and cerebellum were the top regions contributing to shared alterations across the CNVs (54). This overlaps with our findings that highlight the cerebellum in several 1q21.1 distal carriers. Likewise, in our study, for certain 1q21.1del CNV carriers, the caudate and accumbens show significant positive deviations, also overlapping with previous findings (41). The reason the implicated regions we identify might differ slightly from previous studies could be that we focused on individual variations from the norm of the population, rather than comparing the average differences between cases and controls. Moreover, although there is some overlap between studies, our study group represents a set of CNV carriers with a somewhat different profile from those studied previously.

### Towards personalized psychiatry: individualized risk profiles and beyond -

When interpreting the normative model outputs, a common pitfall is to default to a case-control thinking paradigm. This interpretation often categorizes individuals into groups, emphasizing group patterns or seeking group effects instead of individual-level results. While brain deviation score maps can be superimposed to uncover commonalities among subjects with identical pathogenic CNVs, it is not a necessity. In our pursuit of understanding pathogenic CNVs and their effects on cognitive functioning and mental health, it is essential to recognize that we cannot solely rely on aggregated group-level data. While grouping subjects that exhibit similar behavioral phenotypes or possess the same CNV can provide insights into convergence points, this approach overlooks the diversity in the effects of pathogenic CNVs on brain structures and behaviors. For instance, those with a CNV linked to cognitive deficits might range from typical cognitive functioning to severe impairment (47,48,63). Arguably, the starting point should be individual patient risk profiles, including all the known risk factors, environment, genetics, and lifestyle, among others. Once we curate these profiles against a reference population, we can start to understand the implications of pathogenic CNVs at an individual level.

Adopting an “individual patient first” approach reshapes our perspective on psychiatry, emphasizing that no two mental disorders are truly identical. Traditional psychiatric diagnoses have been classified into separate mental disorders, each presumed to have distinct origins and symptomologies. However, we have come to realize that such boundaries are far from clear-cut. Most patients with one diagnosis often have one or more comorbid conditions (22). This wide-ranging clinical manifestation, coupled with multifactorial etiological risk factors and comorbidities, underscores that most psychiatric disorders do not correspond to single disease entities. Initiatives like the Research Domain Criteria (RDoC) (64) have aimed to transition to more dimensional approaches, current categorizations like the DSM-V persist, even amidst challenges such as coexisting conditions and the complexity of categorizing patients. In this paper, we made a starting point toward understanding the individual implications of pathogenic CNVs on several brain phenotypes, cognition, and mental health.

### Addressing the limitations of genetic studies -

One limitation of our study is the lack of longitudinal imaging data for the pathogenic CNV carriers, particularly within the younger age range. While our model covers the entire age range, our conclusions are constrained to participants within the older age range due to the limited availability of CNV data to only the UK biobank. As we know that CNVs play a large role in neurodevelopment (10,41), it is important to track participants with these CNVs longitudinally during the age range where their influence is the largest. Furthermore, we acknowledge that in the study the individual deviation scores might be partly driven by effects outside of the CNVs, for instance, the remaining genome. Research has shown that environmental factors can lead to molecular “scars”, impacting brain function over time and contributing to disorders like schizophrenia (20). For instance, individuals with a 1q21.1 distal deletion or duplication have a higher risk for cognitive impairment, schizophrenia, and micro/macrocephaly (65). Nevertheless, this CNV may also be present in individuals who are otherwise healthy. Therefore, it is likely that the expression of mental disorders associated with this CNV is influenced by a combination of environmental factors and an individual’s lifestyle choices. Epigenetic pathways can offer a valuable perspective for studying these factors. While initial efforts to understand brain-environment-genetic interactions have started (66), an important question remains what the role of CNVs is in this complex picture. Currently, we are developing longitudinal normative models (26,67), which would help us to understand critical time frames and can be used to model the interplay between genetics, environment, and brain function. In future investigations, we aim to integrate the normative modeling framework with these longitudinal samples to examine the dynamics of genetic markers and environmental influences over time.

Another limitation of current normative or reference-based studies is the disproportionate representation of individuals of European ancestry in our reference samples (25,68). We attempted to make this bias explicit in our study by including this information within the demographic tables of our reference samples. However, it remains evident that our reference models are likely not representative of individuals with diverse ancestral backgrounds beyond those in the reference sample. This limitation reduces the generalizability of our models until larger more diverse datasets become available.

In conclusion, addressing these complexities is crucial for advancing our understanding of genetic influences on mental health, requiring a multi-dimensional approach that integrates genetics, environment, and developmental aspects. The present study marks an initial step toward unraveling the impacts of pathogenic CNVs on the brain and cognition at an individualized level.

## Code and data availability -

The data used in this study are all attained from publicly available resources (Cam-CAN: https://www.cam-can.org/index.php?content=dataset; PNC: https://www.nitrc.org/projects/pnc; UKB: https://www.ukbiobank.ac.uk; OASIS: https://www.oasis-brains.org; HCP: https://www.humanconnectome.org/study/hcp-young-adult). The code to create the reference models is available at https://github.com/amarquand/PCNtoolkit. Scripts for creating the visualizations will be available on GitHub (https://github.com/CharFraza).

## Data Availability

All data utilized comes exclusively from open-source neuroimaging databases, which are available at Cam-CAN (https://www.cam-can.org/), HCP (https://www.humanconnectome.org/), Oasis (https://www.oasis-brains.org/), PNC (https://www.med.upenn.edu/bbl/philadelphianeurodevelopmentalcohort.html), and UK Biobank (https://www.ukbiobank.ac.uk/).

## Acknowledgments -

This research was supported by grants from the European Research Council (ERC, grant “MENTALPRECISION” 10100118) and the Dutch Organisation for Scientific Research (VIDI grant 016.156.415). This research has been conducted using the UK Biobank resource under application number 23668. Rune Bøen and Ida Sønderby are supported by the Research Council of Norway (#223273) and the South-Eastern Norway Regional Health Authority (#2020060). In addition, Ida Sønderby is supported by the European Union’s Horizon2020 Research and Innovation Programme (CoMorMent project; Grant #847776) and Kristian Gerhard Jebsen Stiftelsen (SKGJ-MED-021).

## Supplemental figures and tables

**Supplemental Fig. 1.**
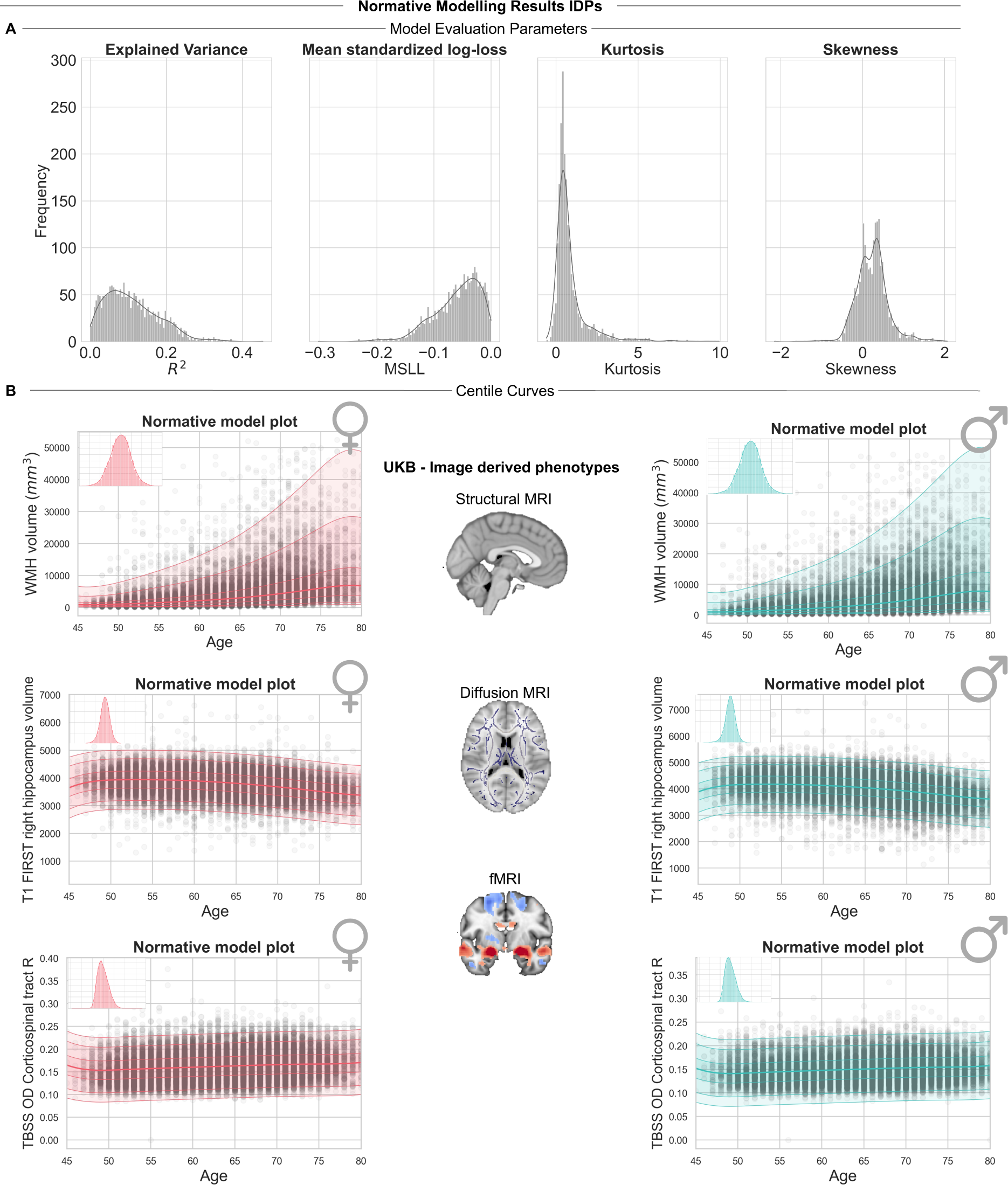
| Overview, normative modeling results IDPs. **A.** Performance metrics on the IDP normative models for the test set. Both skew and kurtosis serve as indicators of the model’s accuracy in estimating shape via warped Bayesian Linear Regression; ideal values approach zero. **B.** Depiction of varied normative trajectories across distinct IDPs.

**Supplemental Fig. 2.**
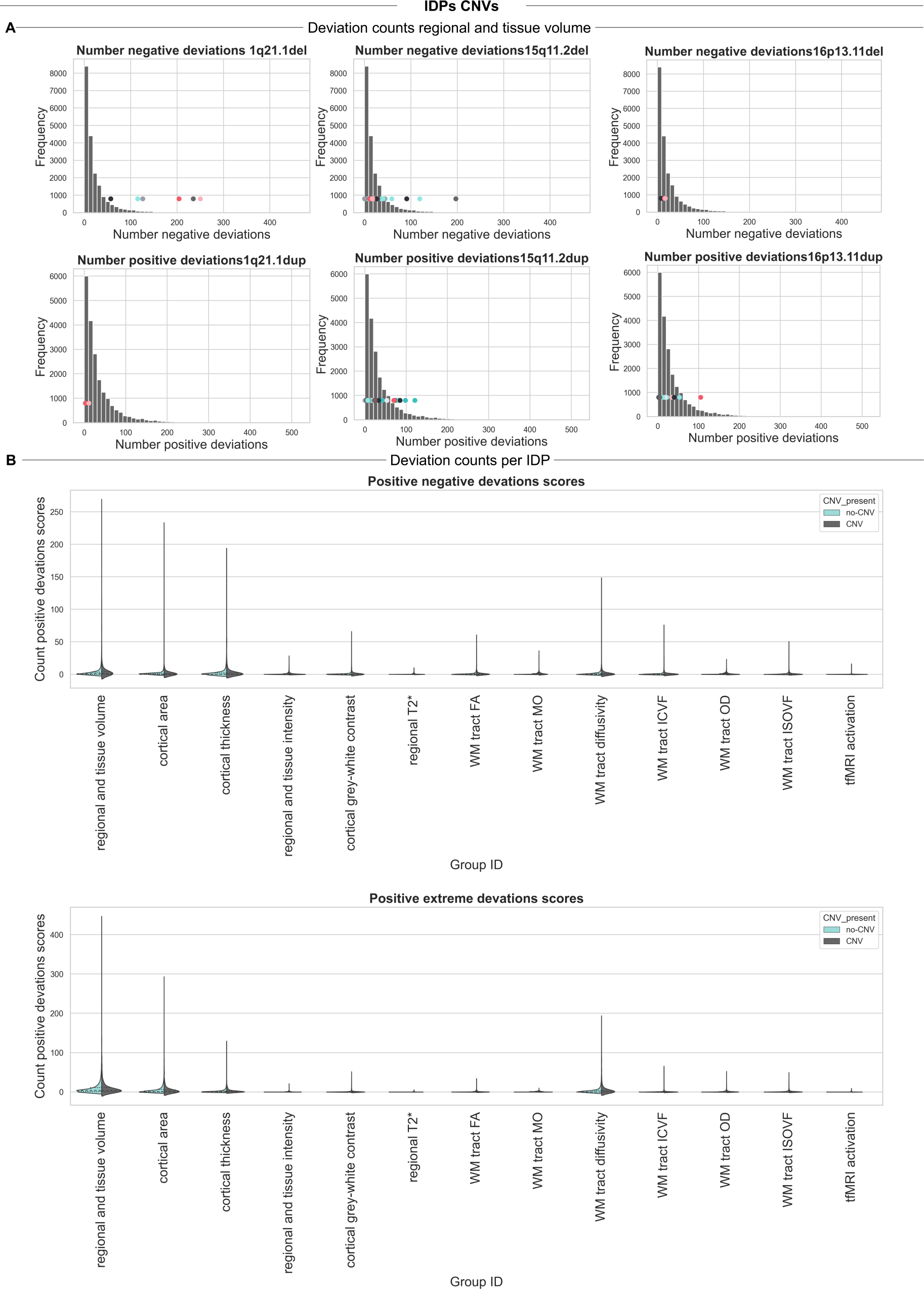
| Deviation counts per IDP category. **A**. Examples of extreme negative deviation counts for the regional and tissue volume IDPs highlighting participants with 1q21.1del, 15q11.2del, 16p13.11del and showing the extreme positive deviation counts for participants with CNV 1q21.1dup, 15q11.2dup, 16p13.11dup. **B**. Showing the positive and negative deviation counts for different IDP categories, comparing participants with or without a CNV.

**Supplemental Fig. 3.**
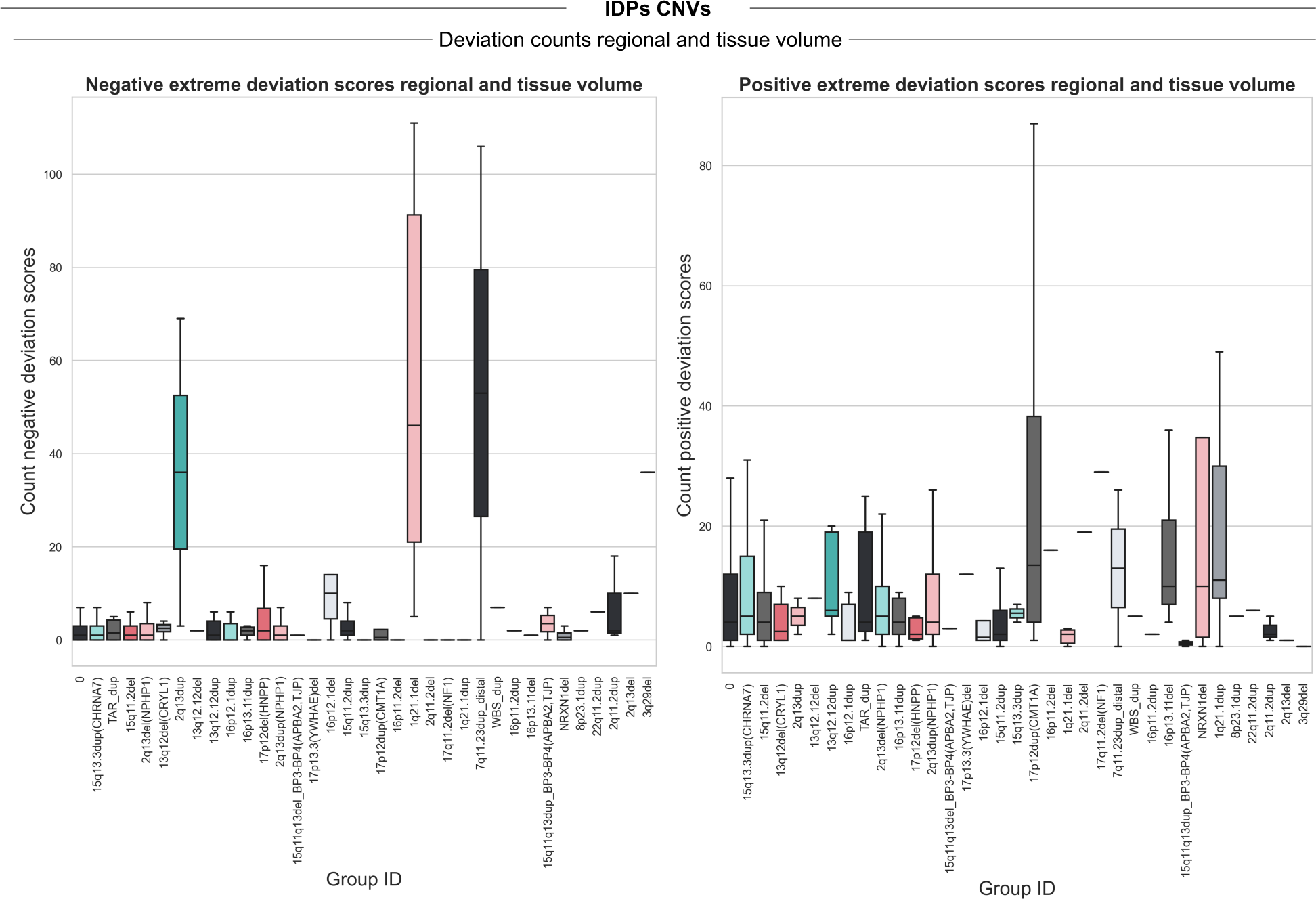
| Overview, deviation counts IDPs per CNV. Showing the positive and negative deviation counts from the IDP normative models for different pathogenic CNVs.

**Supplemental Fig. 4.**
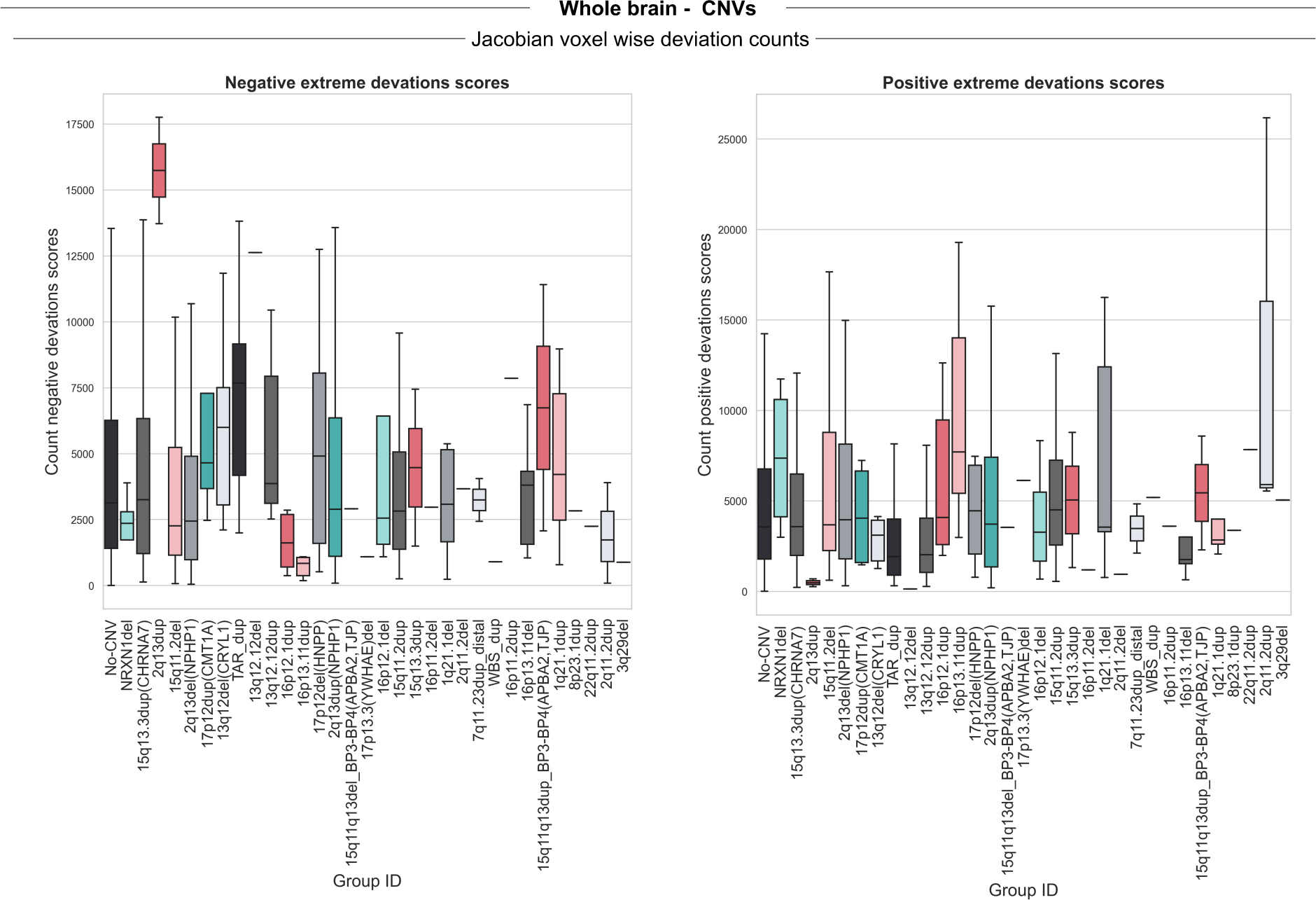
| Overview, deviation counts voxel-wise Jacobian model per CNV. Showing the positive and negative deviation counts from the voxel-wise Jacobian normative models for different pathogenic CNVs.

**Supplemental Fig. 5.**
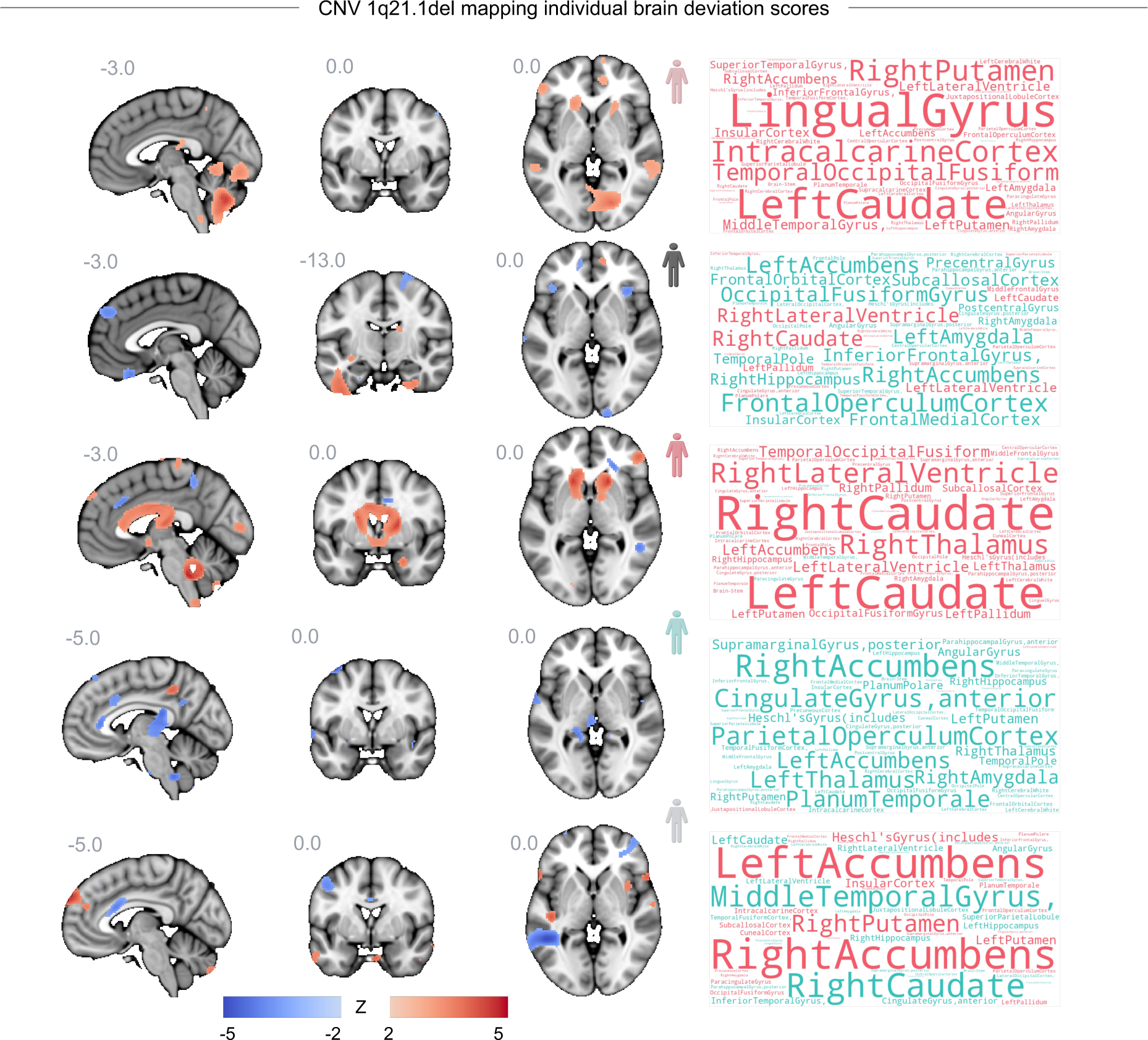
| Individual brain map and deviation scores. Showing the individual positive and negative deviation scores for all five participants with a 1q21.1 distal deletion, and the accompanying word clouds, indicating the mean z-value for each brain area, using the Harvard cortical and subcortical atlas.

**Supplemental Fig. 6.**
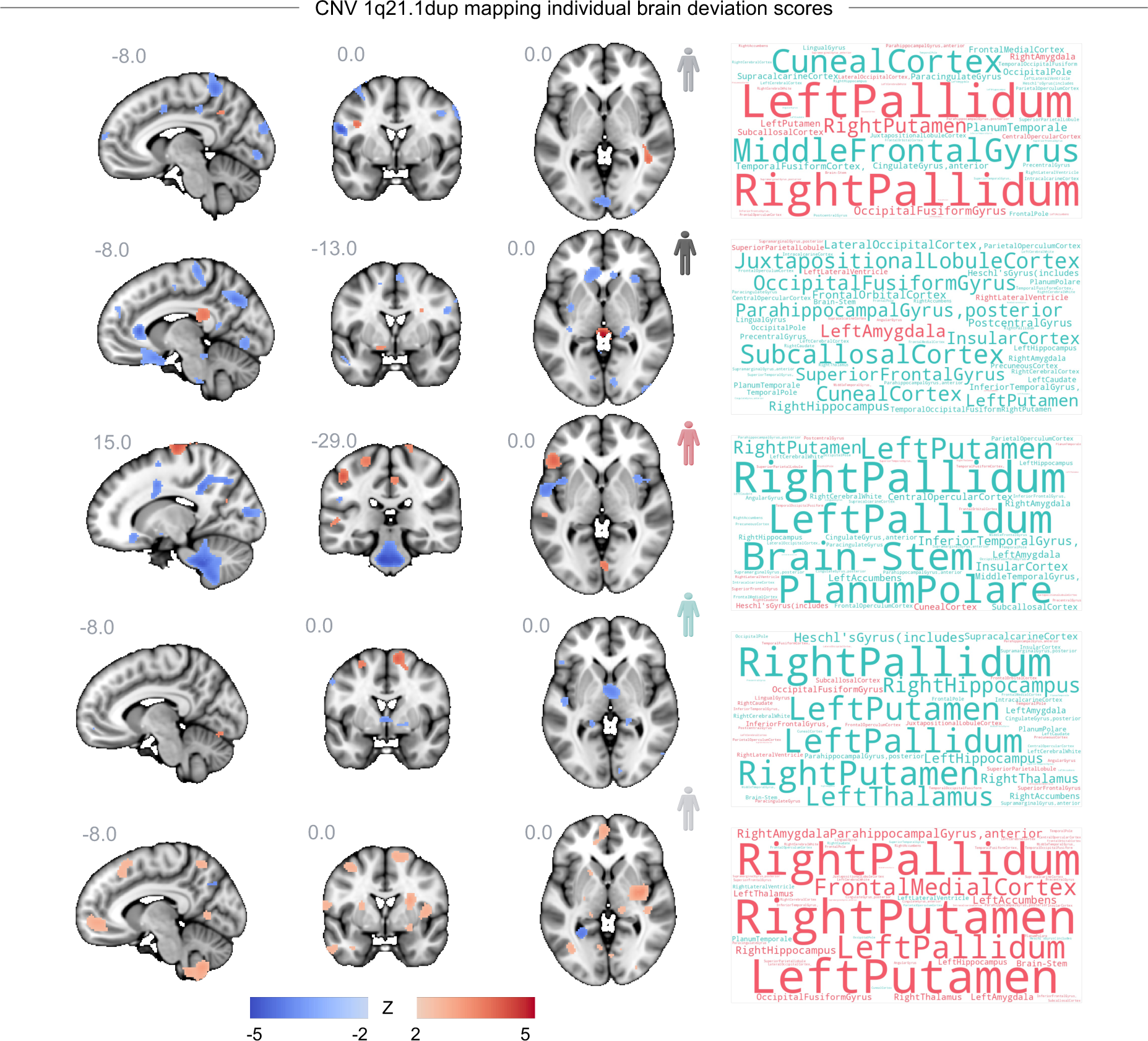
| Individual brain map and deviation scores. Showing the individual positive and negative deviation scores for all five participants with a 1q21.1 distal duplication, and the accompanying word clouds, indicating the mean z-value for each brain area, using the Harvard cortical and subcortical atlas.

**Supplemental Fig. 7.**
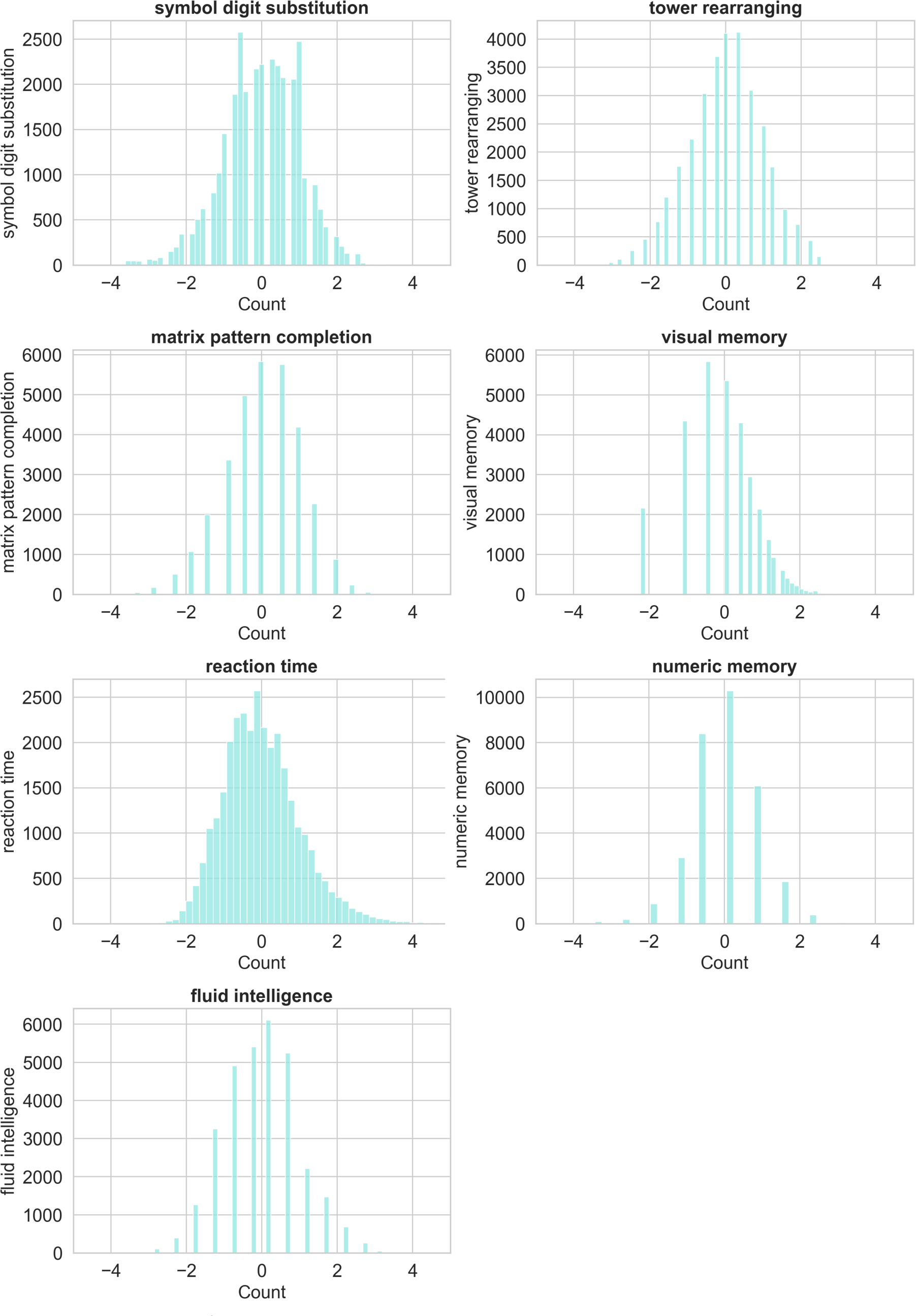
| Overview cognitive tests UK Biobank. Showing the histograms of the cognitive tests included in the study to create a general cognitive component using a principal component analysis and selecting the first principal component.

**Supplemental Fig. 8.**
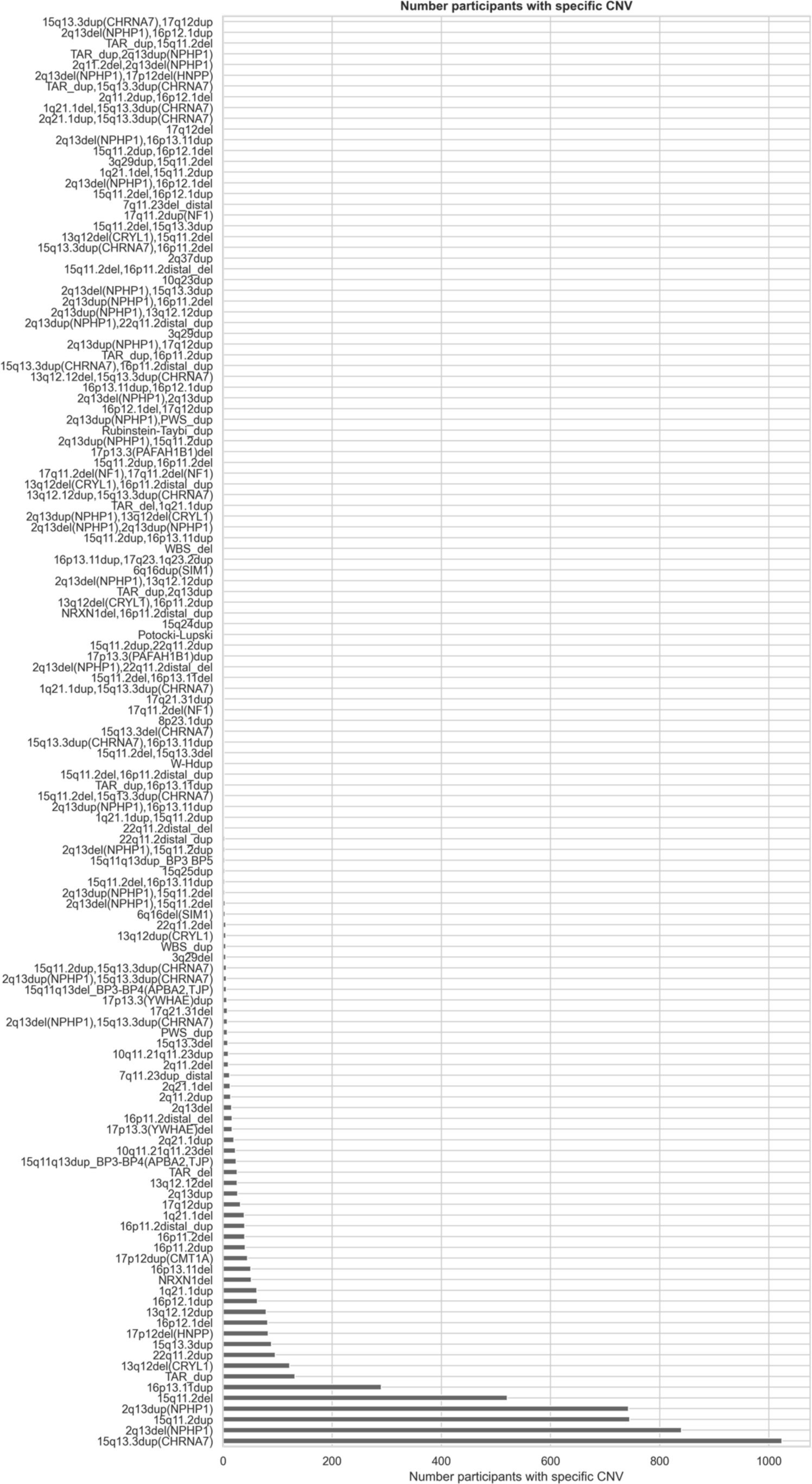
| Overview 92 pathogenic CNVs present in the UK Biobank.

**Supplemental Fig. 9.**
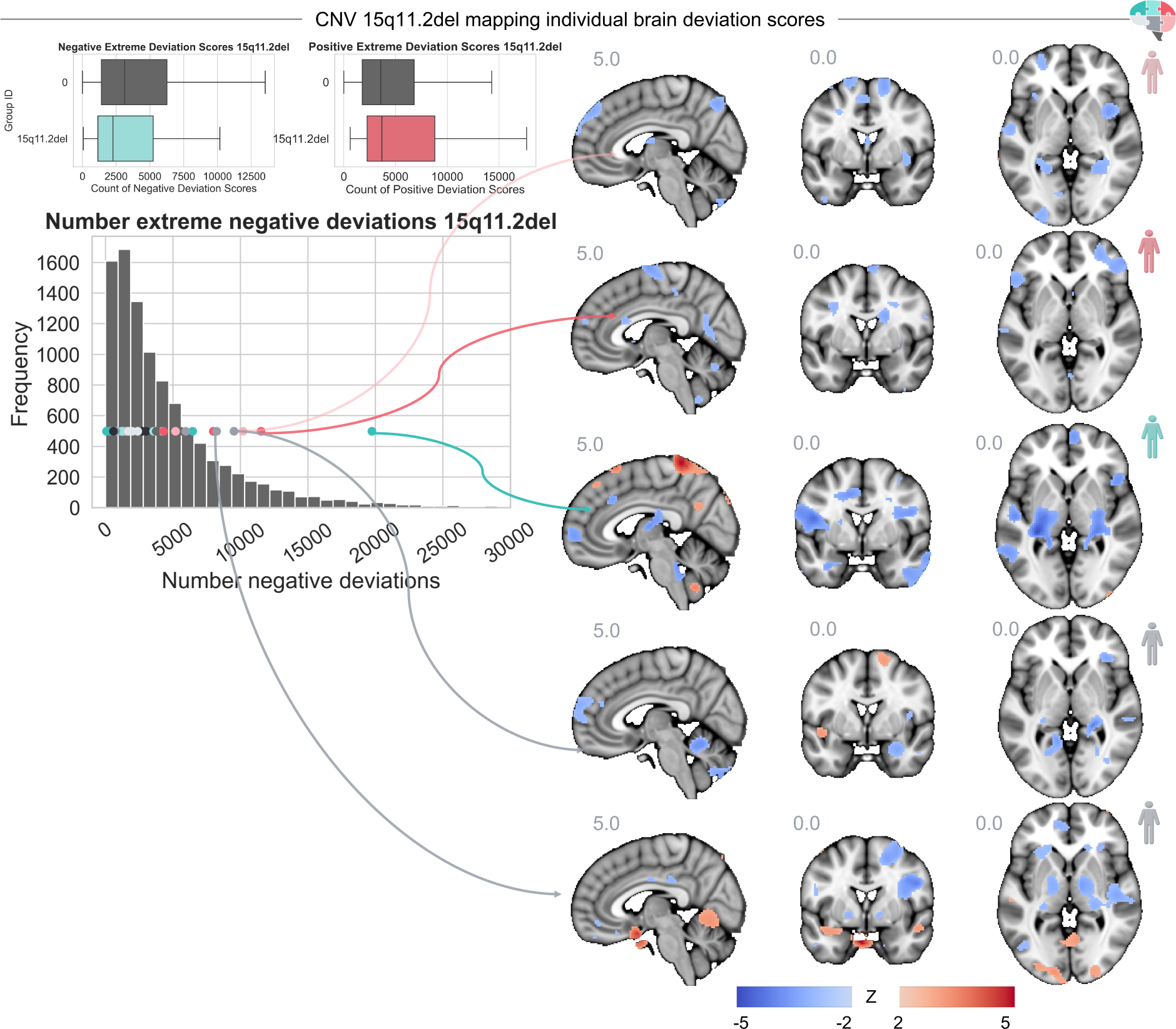
| Individual risk profiles 15q11.2 deletion. Showing the counts of extreme positive and negative deviation scores (|Z|>2) among participants with a 15q11.2 deletion in contrast to participants without a pathogenic CNV. Left: Dots show each individual 15q11.2 deletion carrier’s position in the distribution. Right: Displaying the profile of five selected 15q11.2 deletion CNV carriers.

**Supplemental Fig. 10.**
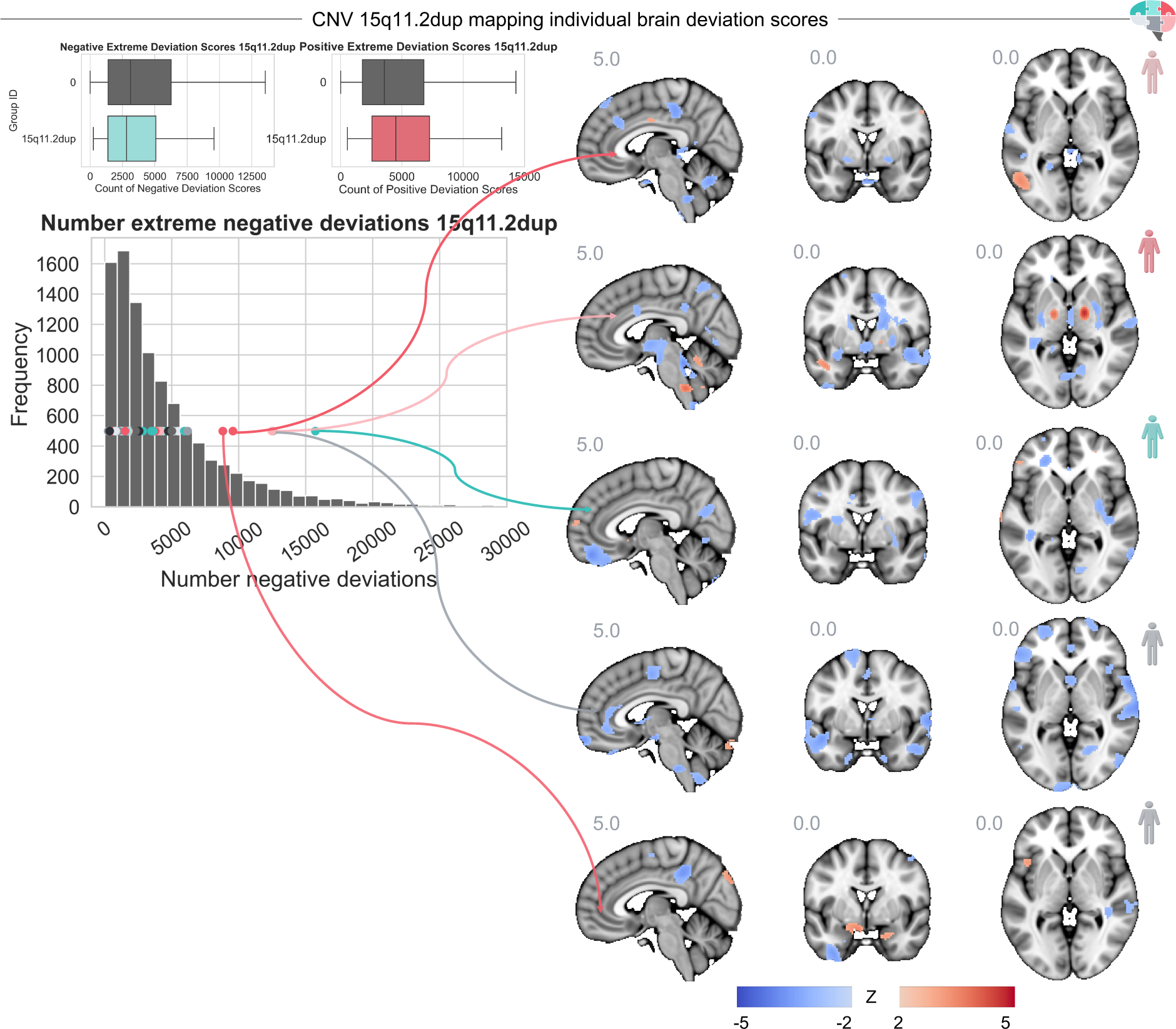
| Individual risk profiles 15q11.2 duplication. Showing the counts of extreme positive and negative deviation scores (|Z|>2) among participants with a 15q11.2 duplication in contrast to participants without a pathogenic CNV. Left: Dots show each individual 15q11.2 duplication carrier’s position in the distribution. Right: Displaying the profile of five selected 15q11.2 duplication CNV carriers.

**Supplemental Fig. 11.**
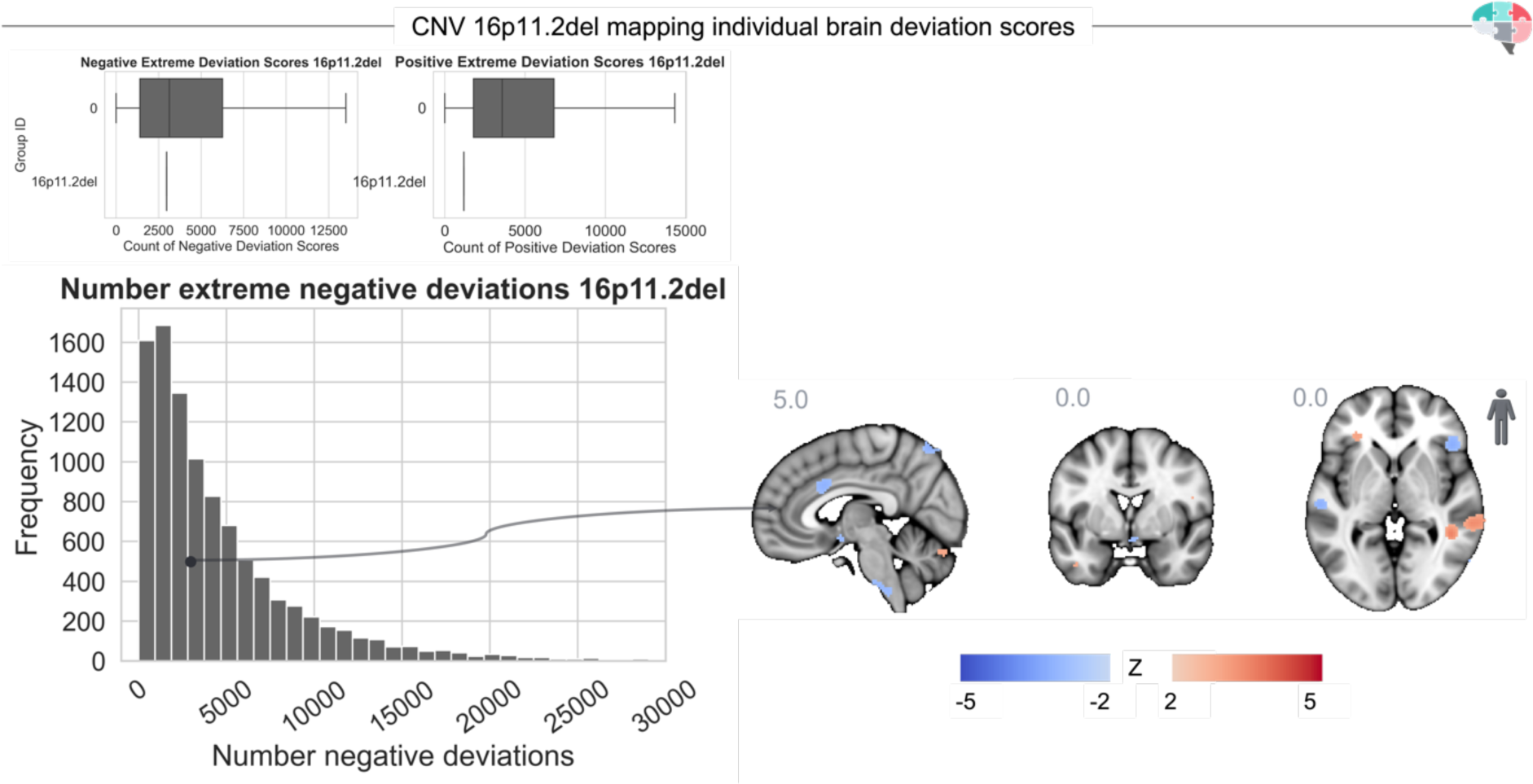
| Individual risk profile 16p11.2 deletion. Showing the counts of extreme positive and negative deviation scores (|Z|>2) among participants with a 16p11.2 deletion in contrast to participants without a pathogenic CNV. Left: Dots show each individual 16p11.2 deletion carrier’s position in the distribution. Right: Displaying the profile of one 16p11.2 deletion CNV carrier.

**Supplemental Fig. 12.**
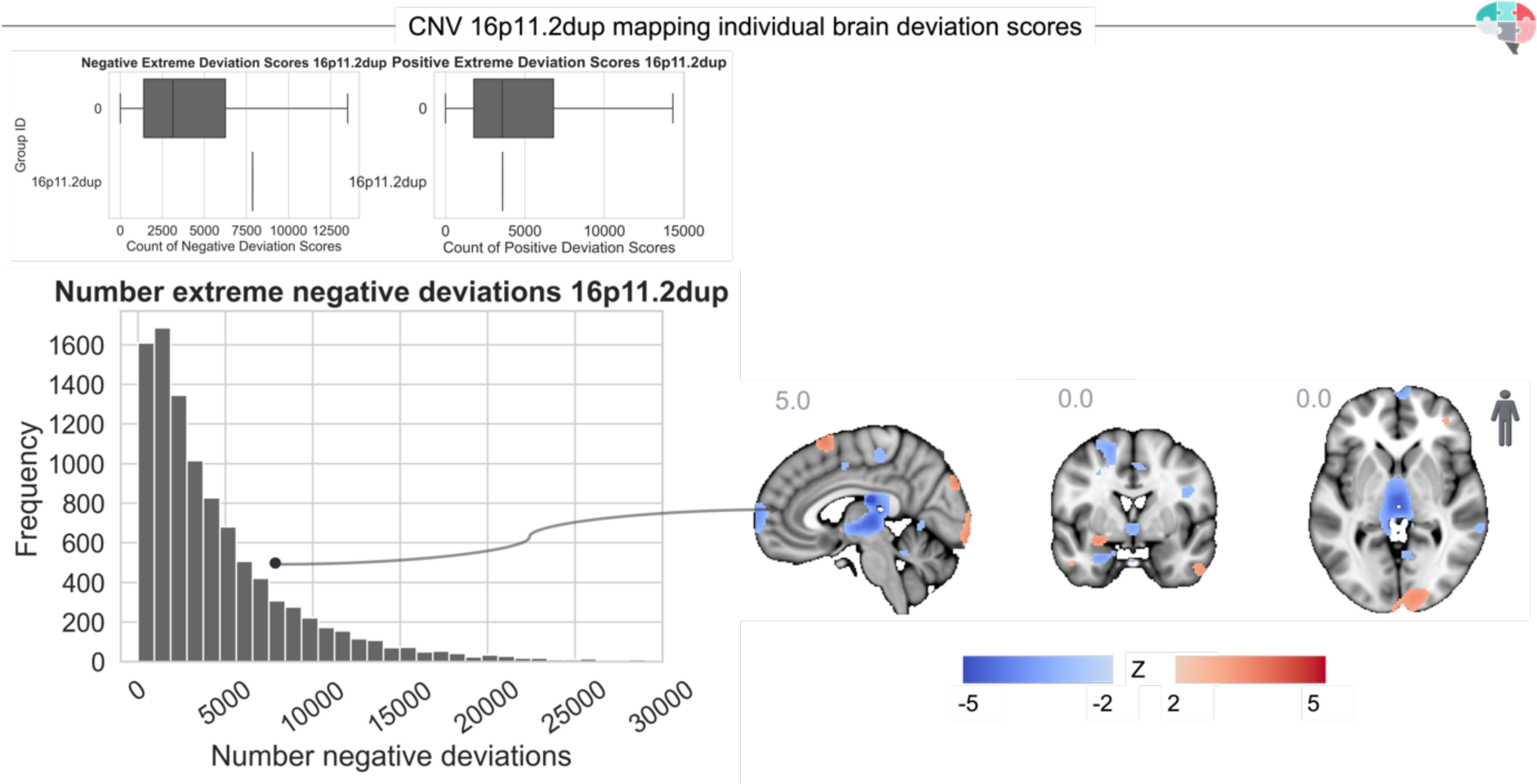
| Individual risk profile 16p11.2 duplication. Showing the counts of extreme positive and negative deviation scores (|Z|>2) among participants with a 16p11.2 duplication in contrast to participants without a pathogenic CNV. Left: Dots show each individual 16p11.2 duplication carrier’s position in the distribution. Right: Displaying the profile of one 16p11.2 duplication CNV carrier.

**Supplemental Fig. 13.**
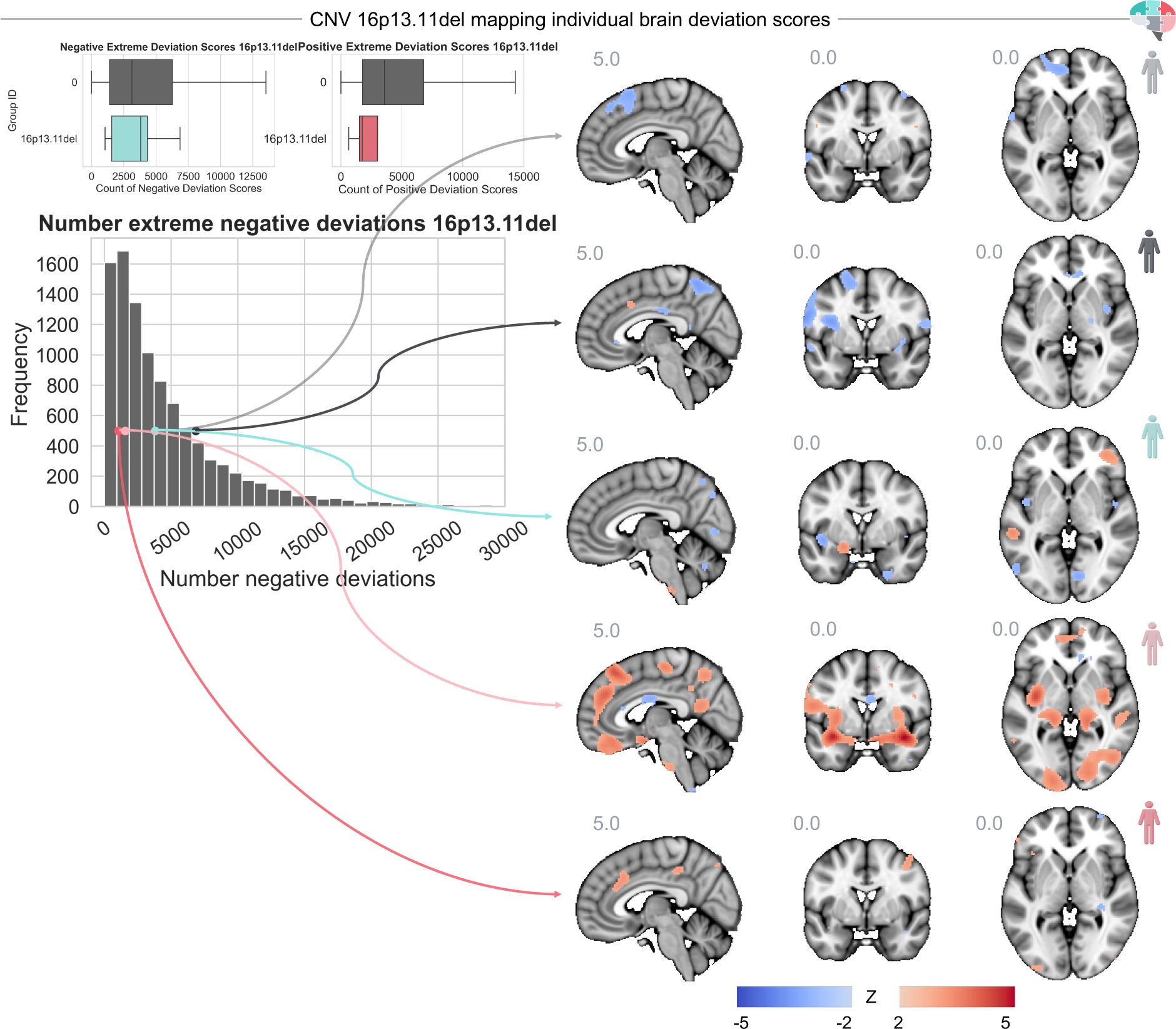
| Individual risk profile 16p13.11 deletion. Showing the counts of extreme positive and negative deviation scores (|Z|>2) among participants with a 16p13.11 deletion in contrast to participants without a pathogenic CNV. Left: Dots show each individual 16p13.11 deletion carrier’s position in the distribution. Right: Displaying the profile of five selected 16p13.11 deletion CNV carriers.

**Supplemental Fig. 14.**
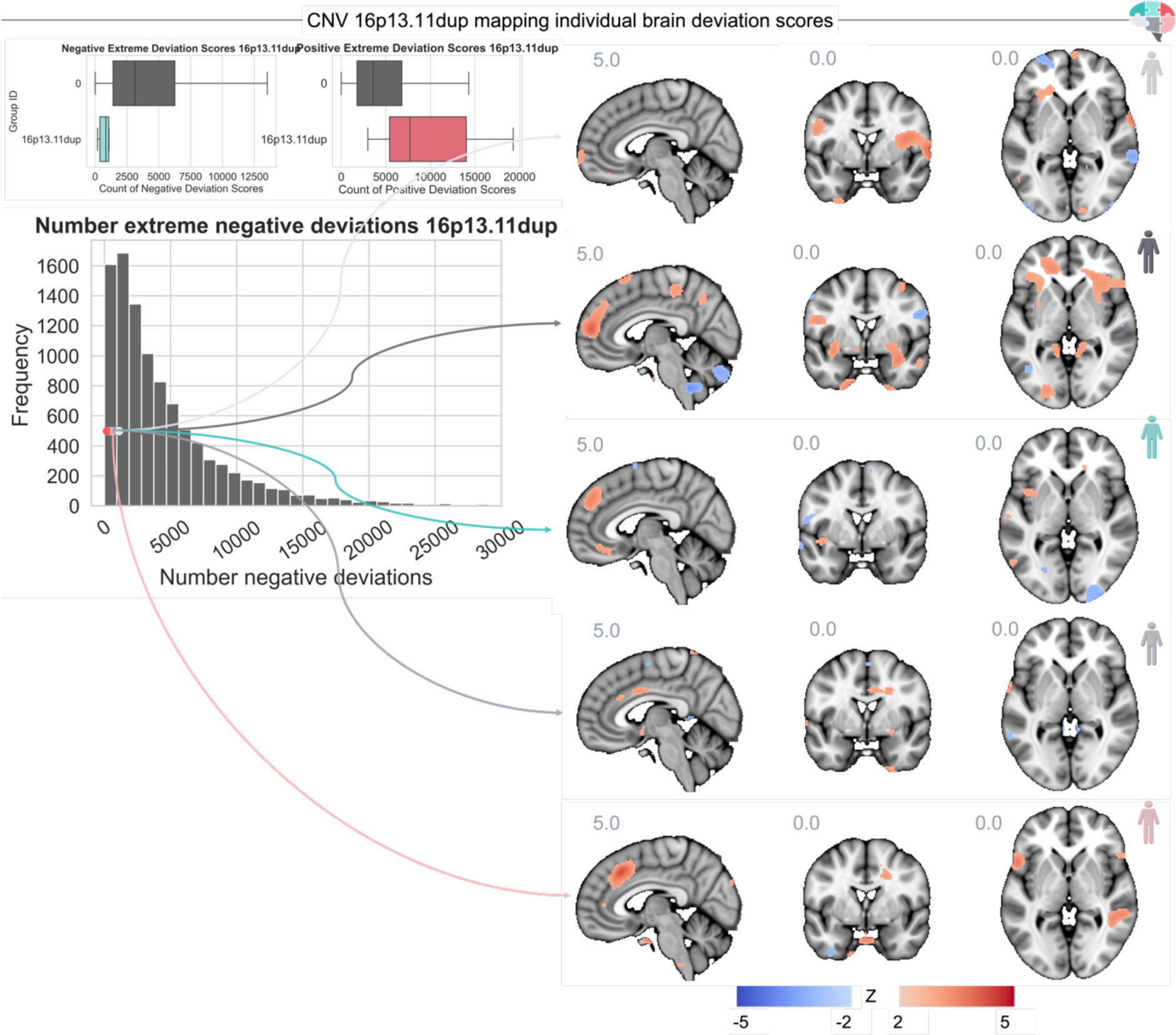
| Individual risk profile 16p13.11 duplication. Showing the counts of extreme positive and negative deviation scores (|Z|>2) among participants with a 16p13.11 duplication in contrast to participants without a pathogenic CNV. Left: Dots show each individual 16p13.11 duplication carrier’s position in the distribution. Right: Displaying the profile of five selected 16p13.11 deletion CNV carriers.

**Supplemental Fig. 15.**
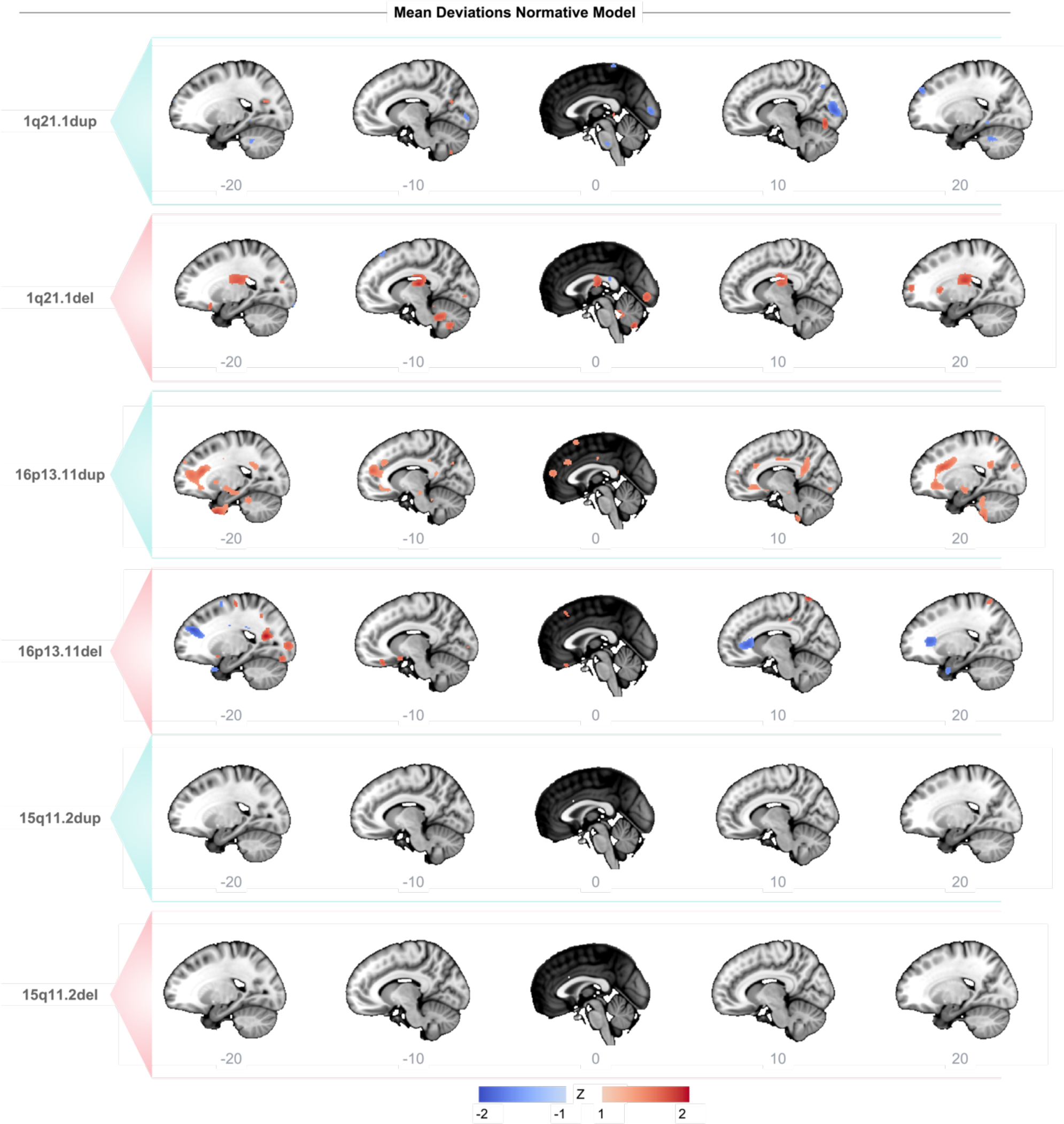
|Convergence of positive or negative deviation scores. The overlapping brain deviation score maps for participants with the 1q21.1 distal, 16p13.11, or 15q11.2del CNVs. On the x-axis, we show different sagittal slices with steps of 10.

**Supplemental Fig. 16.**
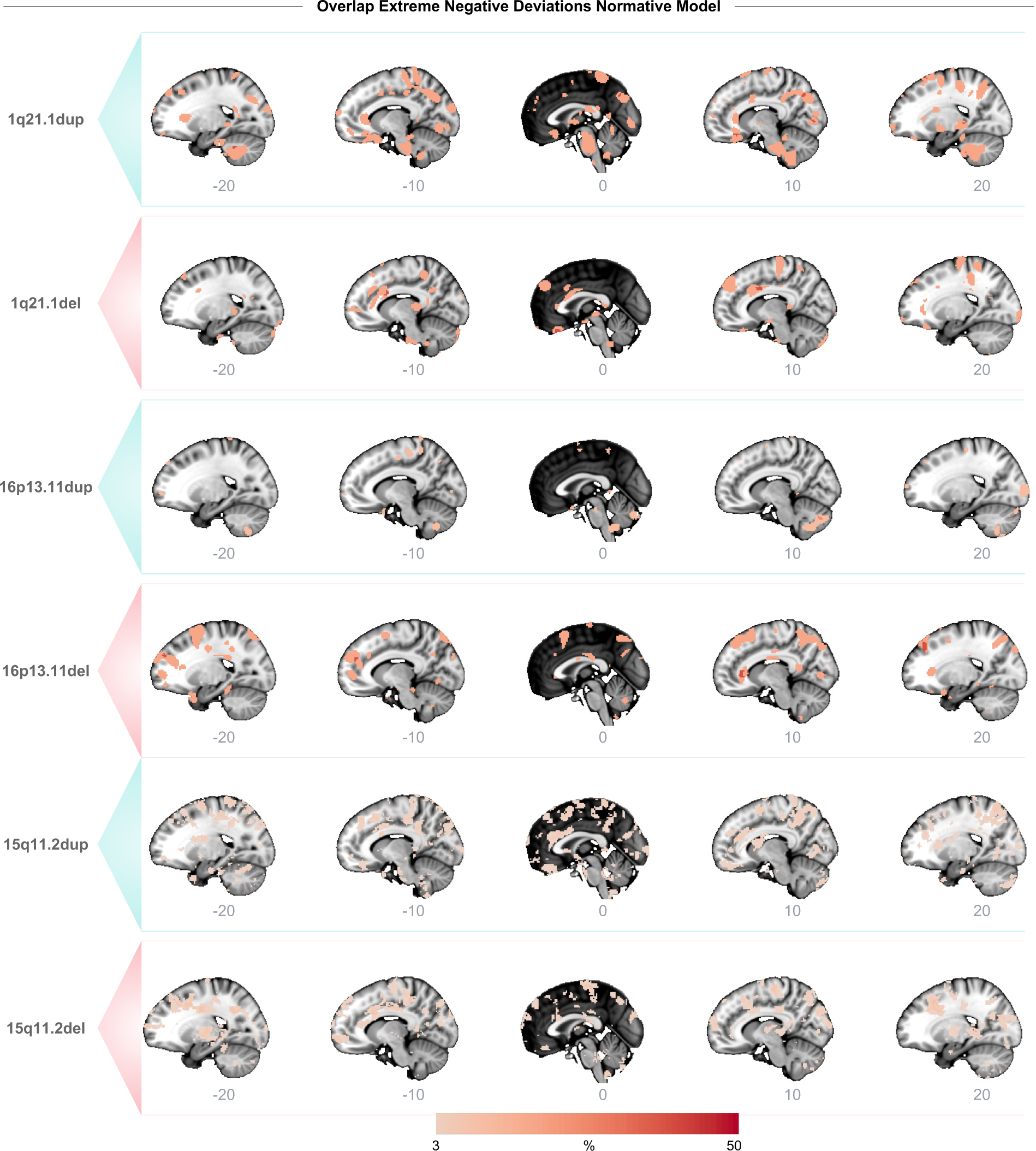
|Percentage overlap of extreme negative deviation scores. Showing the percentage of overlapping subjects with extreme negative brain deviation scores (Z<-2) for participants with the 1q21.1 distal, 16p13.11, or 15q11.2del CNVs. On the x-axis, we show different sagittal slices with steps of 10.

**Supplemental Fig. 17.**
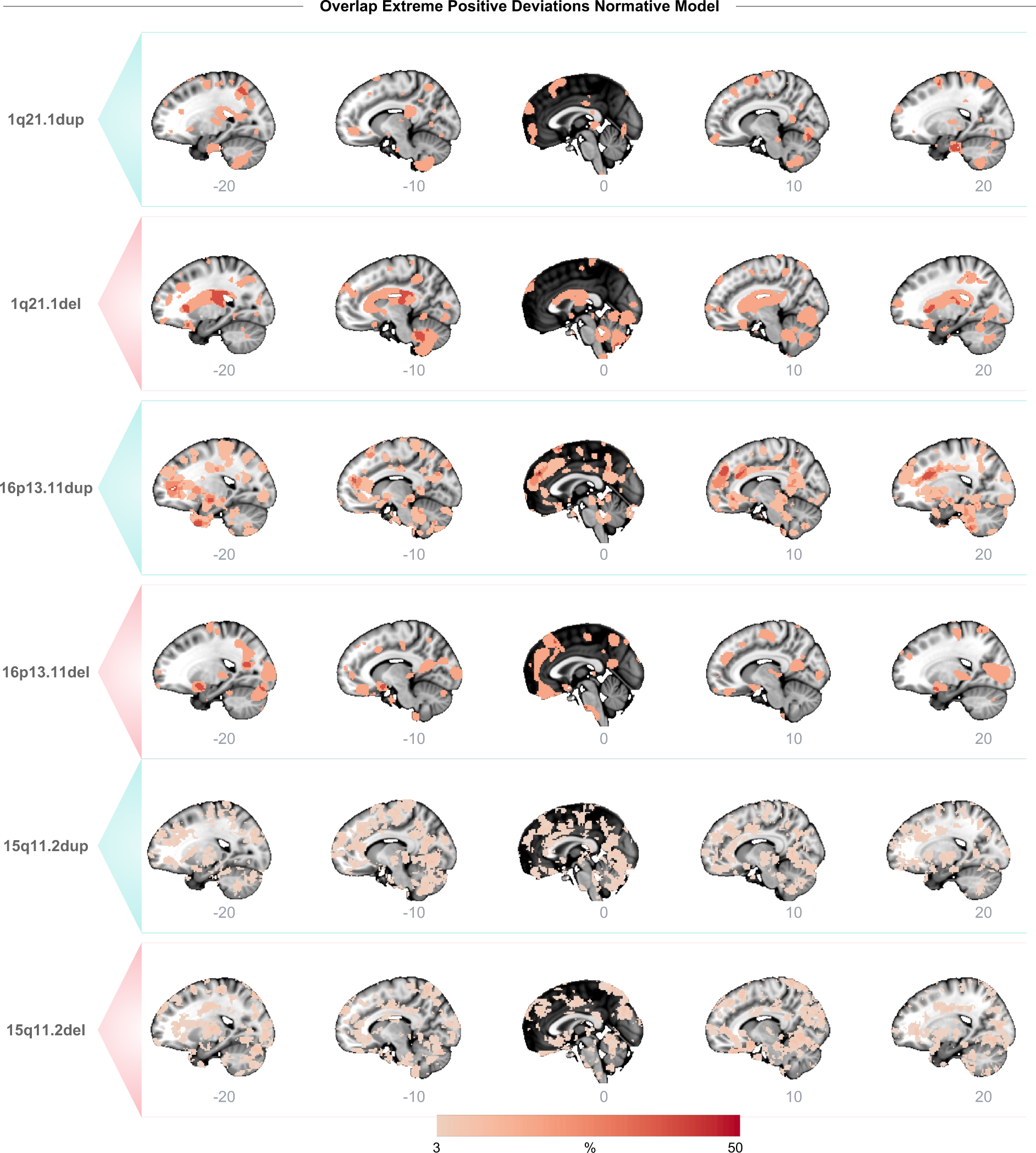
|Percentage overlap of extreme positive deviation scores. Showing the percentage of overlapping subjects with extreme negative brain deviation scores (Z>2) for participants with the 1q21.1 distal, 16p13.11, or 15q11.2del CNVs. On the x-axis, we show different sagittal slices with steps of 10.

**Supplemental Fig. 18.**
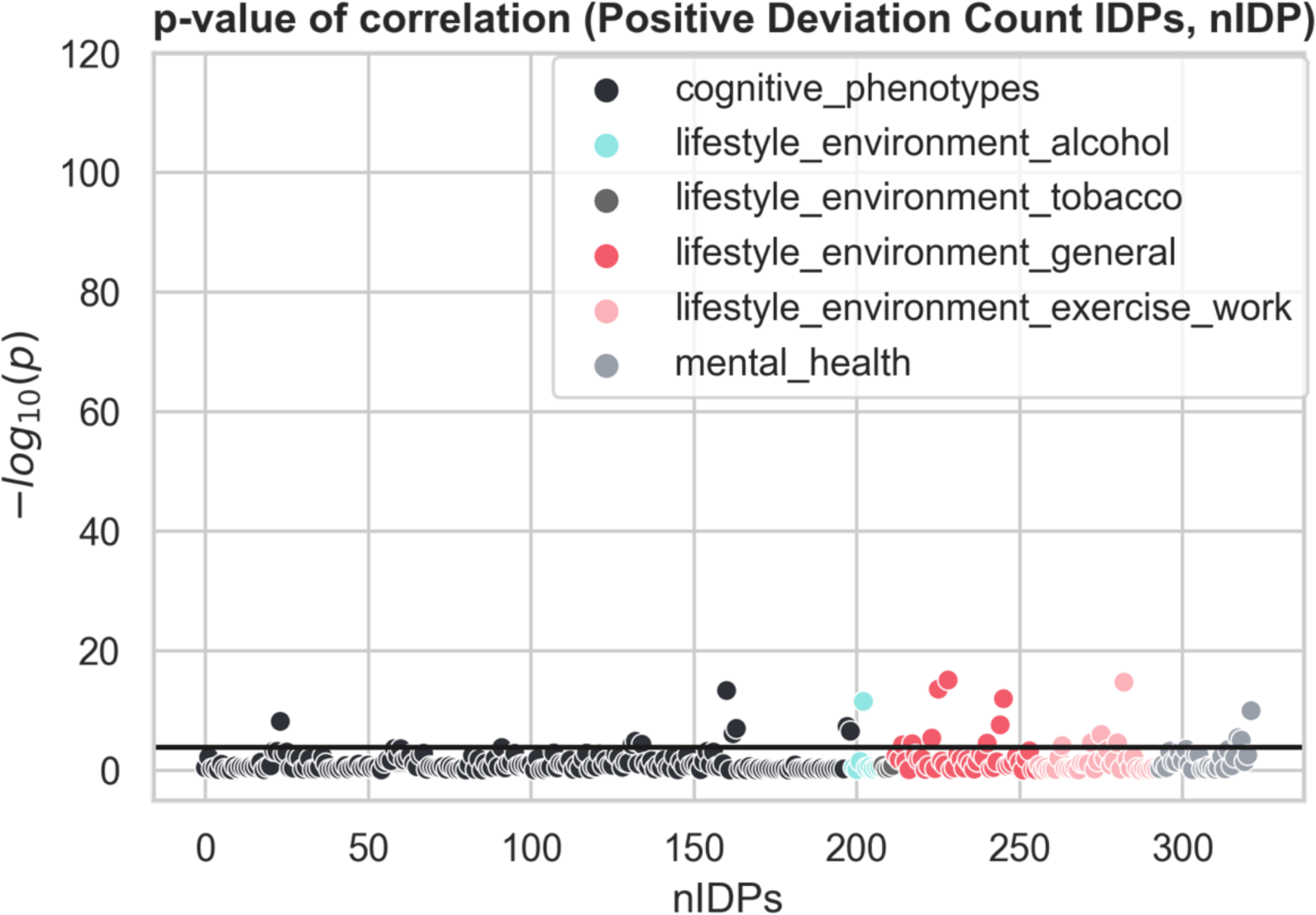
|Manhattan plot spearman correlation nIDPs and extreme positive deviation count IDPs. The black line indicates the p-value threshold Bonferroni corrected. The total number of correlations passing the p-value threshold: 28.

**Supplemental Fig. 19.**
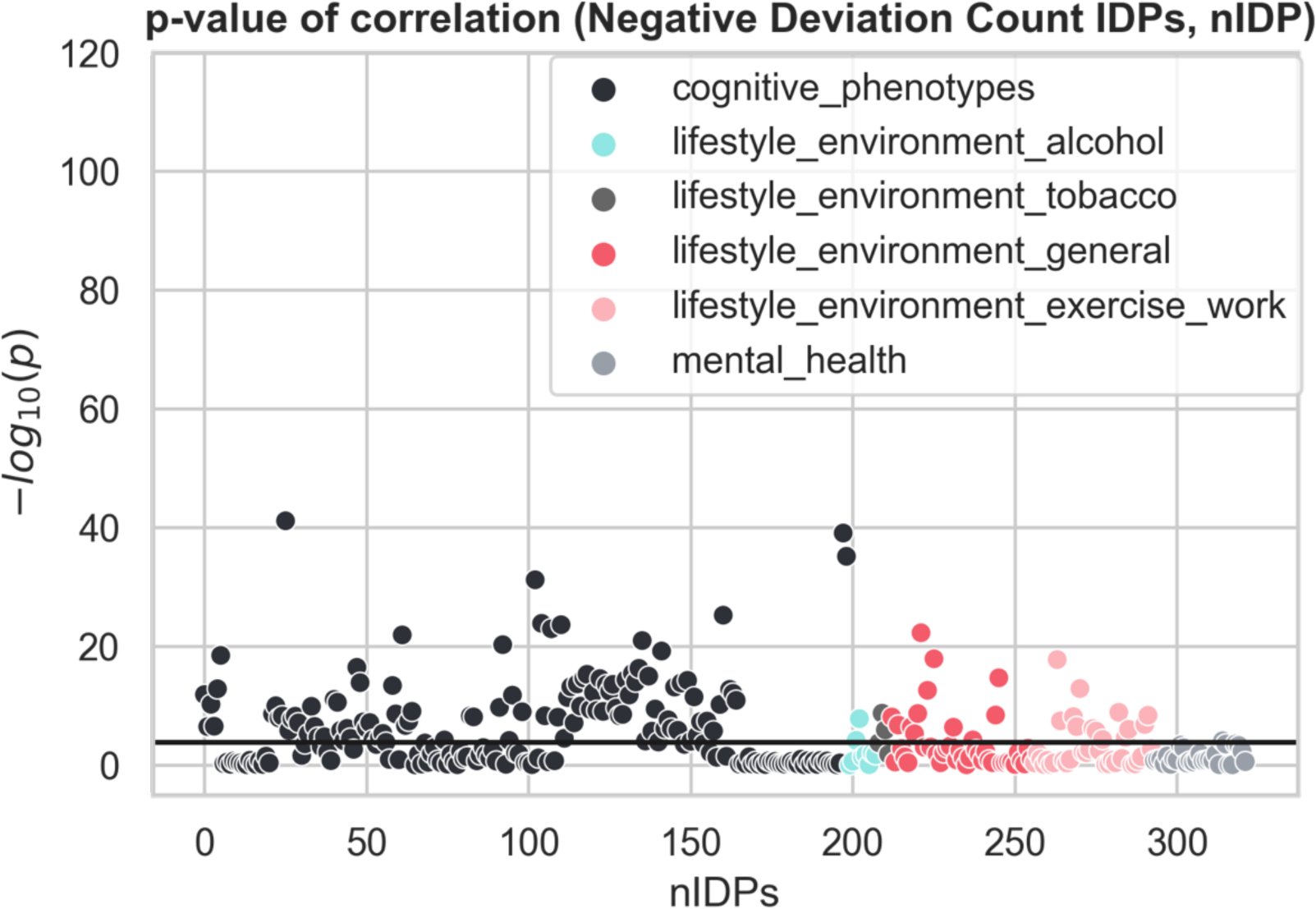
|Manhattan plot spearman correlation nIDPs and extreme negative deviation count IDPs. The black line indicates the p-value threshold Bonferroni corrected. The total number of correlations passing the p-value threshold: 137.

**Supplemental Fig. 20.**
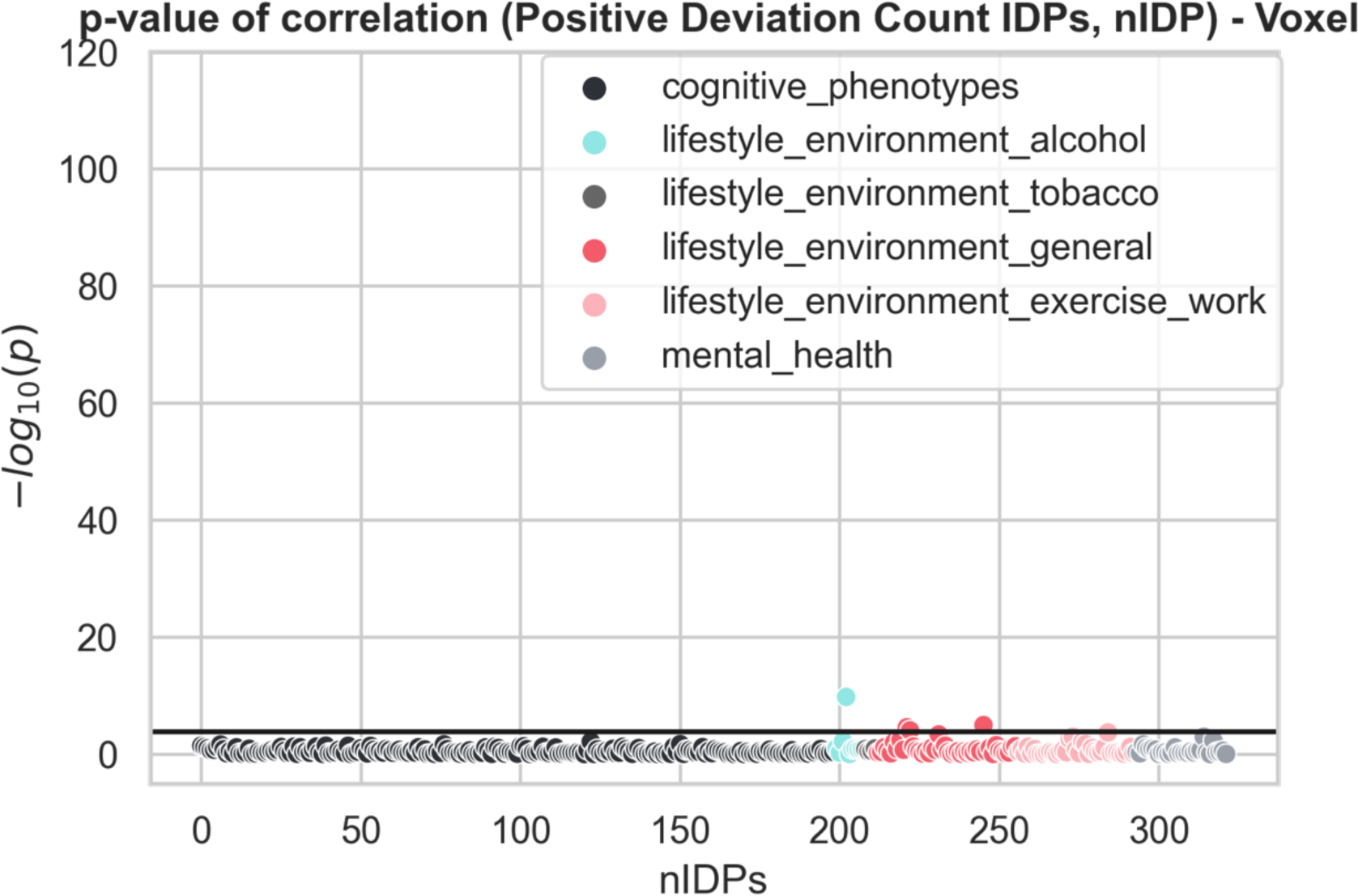
|Manhattan plot spearman correlation nIDPs and extreme positive deviation count Jacobian voxels. The black line indicates the p-value threshold Bonferroni corrected. The total number of correlations passing the p-value threshold: 4.

**Supplemental Fig. 21.**
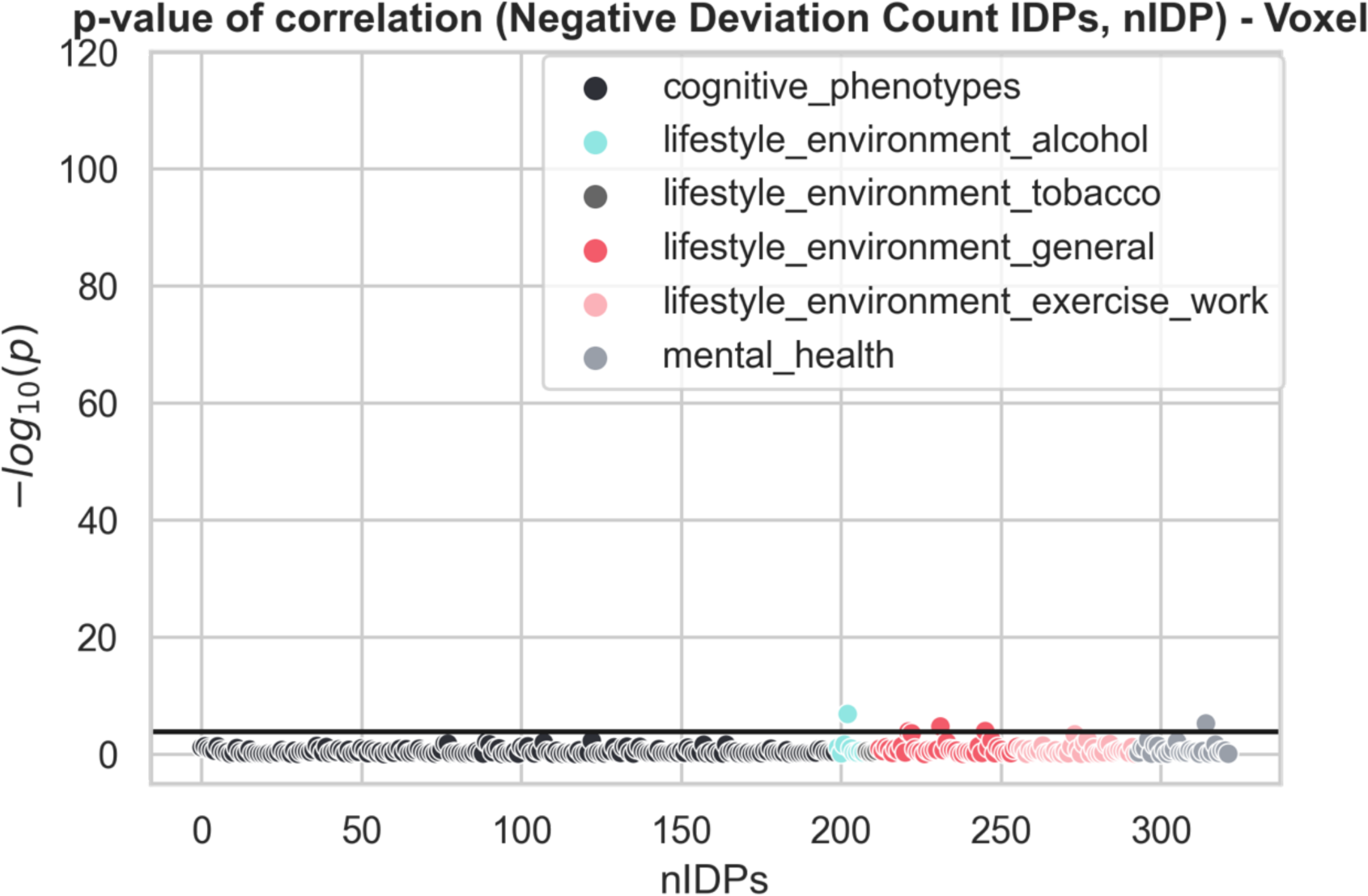
|Manhattan plot spearman correlation nIDPs and extreme negative deviation count Jacobian voxels. The black line indicates the p-value threshold Bonferroni corrected. The total number of correlations passing the p-value threshold: 5.

## Reference cohort assembly

### Data – Image-derived phenotypes

In this study, two distinct types of analyses were performed using different datasets. For the first analysis, a dataset containing the image-derived phenotypes (IDPs) was used. In alignment with prior research, these IDPs underwent preprocessing using FUNPACK (39), an automated toolkit for normalization, parsing, and cleaning, developed at the Wellcome Centre for Integrative Neuroimaging. The set of IDPs comprises three primary imaging modalities: structural, functional, and diffusion MR data. Encompassed within these IDPs are diverse metrics, ranging from global measurements like total brain volume to more intricate assessments such as interregional connectivity within the brain. In total, the data of 44,456 participants from the UK biobank and 2084 IDPs were used in the analysis. For a detailed description of all the IDPs and the quality control performed within the UK biobank readers can refer to (38).

### Data - Whole brain voxel-wise Jacobian model

The subsequent analysis involved constructing a whole-brain voxel-wise model, which entailed combining the T1-weighted anatomical images from seven distinct sites to form a comprehensive normative sample. A detailed breakdown of each site’s demographics, including factors such as site, gender, and age, can be found in Supplementary Table 1. All the data employed in this study were sourced from publicly accessible repositories: Cam-CAN (34), HCP (35), Oasis (36), PNC (37), and UK Biobank (38). For the UK Biobank, exclusively data from the initial imaging visit were included. Visual inspection was performed to ensure data quality (24), resulting in a total of 19,620 participants with MRI scans passing the quality control assessment. The data were preprocessed with a registration to a standard spatial reference using FSL’s FLIRT and FNIRT, with the commands cmd_fsl_anat and cmd_add_affine to perform anatomical preprocessing and affine transformation to MNI space, for further detail see (69). Within the framework of a voxel-based model, we opted for the Jacobian determinants (T1_to_MNI_aff+nonlin_jac.nii.gz images). Jacobian determinant images derived from the non-linear image registration to the MNI152 space indicate the degree of local volume expansion or contraction in various regions of the brain when compared to a standard reference brain template (MNI152 space). These images are used in neuroimaging to analyze and visualize how brain structures change in size or shape in relation to the chosen reference template. A Jacobian determinant measures the local expansion or contraction factor at each voxel in the brain. Values higher than one indicate volume expansion in that region, while values below one indicate volume contraction. The magnitude of the value can also provide information about the degree of change. This approach removes the need for arbitrary distinctions between white and gray matter and has exhibited robust correlations with specific demographic variables (40).

### CNVs UK Biobank

We identified CNVs based on the returned dataset from Crawford et al. (49). For details on the preprocessing and calling pipeline and quality control of the CNVs see (47,48). Briefly, their quality control included genotypic call rate <0.96, waviness factor of <−0.03 and >0.03, >30 CNVs per person, and log R ratio s.d. of >0.35. Among the participants with IDPs data present 263 individuals had CNVs. Among the participants with quality-controlled Jacobian neuroimaging data, 375 individuals had a pathogenic CNV.

### Normative model formulation

We employed a Bayesian linear regression model (BLR) with likelihood warping for the normative analysis. For an in-depth explanation of the mathematical framework, refer to (43), here we will briefly paraphrase the method. Python version 3.8 and PCNtoolkit version 0.28 were used for all statistical analyses. Our dataset was divided into a 50-50 train-test split, with the test set including all participants with pathogenic CNVs. Covariates encompassed age, binary gender, and binary site ID within the covariance matrix. Additional variables could be incorporated into the model, altering the definition and interpretation of the z-scores. For instance, introducing a global measure such as total intracranial volume (TIV) as a covariate would necessitate understanding the local z-scores relative to this additional factor. Morphometric variation models were estimated using Jacobian determinants from non-linear registration, utilizing the warped BLR algorithm from PCNtoolkit. This algorithm employed Sinarcsinh likelihood warping and Powell optimization. To concisely present the method: we take ***Y*** = (*y_nd_*) ∈ ℝ*^N^*^×*D*^ with *y_nd_* the *d*-th neuroimaging variable, voxel or IDP, of the *n*-th subject. We collected the covariates into one matrix ***X*** = (*x_nm_*) ∈ ℝ*^N^*^×*M*^, where *x_nm_* is the *m*-th covariate of the *n*-th subject. To keep the notation concise we will concentrate on one variable, labelled *d* and drop the subscript. Thus, for every IDP we denote ***y*** = (*y*_1_, …, *y_N_*)*^T^* and take the set of independent variables ***x**_n_*, = (*x_n1_*, …, *x_nm_*)*^T^*. For every subject, we specified the model as follows:

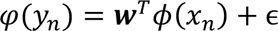

Where, **w**^T^is the estimated vector of weights, ϕ(**x**) a basis expansion of the covariate vector *x_n_* In our case, a cubic B-spline basis expansion with 5 evenly spaced knots was chosen. Empirically, this was enough to capture the curvature in space caused by the age covariate. ɛ_s_ = 𝒩(0, β^-1^) is a Gaussian noise distribution with mean zero and noise precision term β (the reciprocal of the variance). The likelihood warping *φ*, a Sinarcsinh function, accommodates non-Gaussianity:

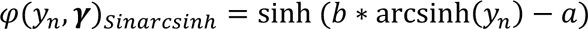

With ***γ*** = (*a*, *b*) the identified parameters for the warping function. We captured the site variation using a fixed-effects model, according to^21,39^. Powell’s conjugate direction method minimized the negative log-likelihood during optimization. Subsequently, z-scores in the warped space were calculated as:

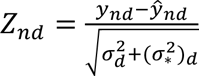

Where, y_nd_ is the true response, ŷ_nd_ is the predicted mean, 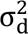 is the estimated noise variance (reflecting uncertainty in the data), and (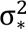)_d_ is the variance attributed to modeling uncertainty, for the full derivations see^63^. Model evaluation employed criteria including R², mean squared log-loss (MSLL), skewness, and kurtosis, providing insights into central tendency, variance, warping function performance, and overall fit.

### Cognitive phenotype prediction

We used the cognitive data extracted from the UK Biobank dataset to explore associations between cognitive attributes and deviations from the normative model. These phenotypes come from seven cognitive tests available in the UK Biobank, as shown in Supplementary Fig. 7. These tests were conducted via a touchscreen questionnaire and encompassed dimensions such as numerical memory, reaction time, fluid intelligence, visual memory, prospective memory, executive function, declarative memory, and non-verbal reasoning (71). Further details about these cognitive assessments in the UK Biobank can be found in (72). To reduce the complexity of cognitive tests while retaining essential information, we employed principal component analysis (PCA) on the cognitive measurements. This approach helps capture a latent factor often associated with overall cognitive ability or the ‘g-factor’ (55). By relating this general cognitive ability to extreme z-deviations (|Z|>2) using Spearman’s rank correlation, we investigated potential associations between cognitive performance and deviations from the norm.

### Estimation of the polygenic scores

The PGS scores were computed for seven major psychiatric disorders using the CS-auto approach (58) on a sample of 377,663 UK Biobank participants of European ancestry. Each PGS was derived from summary statistics of recent Genome-Wide Association Studies (GWAS) and standardized for the subsample used in this study. An overview of the discovery of GWAS studies is presented in Table 1. For further details of the PGS calculation readers can refer to the manuscript (59).

PGSs were computed for the following psychiatric disorders:

1. Attention-Deficit/Hyperactivity Disorder (PGS-ADHD) (73)
2. Autism Spectrum Disorder (PGS-ASD) (74)
3. Major Depressive Disorder (PGS-MDD) (75)
4. Anxiety Disorders (PGS-ANX) (76,77)
5. Schizophrenia (PGS-SCZ) (78)
6. Bipolar Disorder (PGS-BIP) (79)
7. Cannabis Use Disorder (PGS-CUD) (80)

### Correlation of the polygenic scores with the deviation scores

18,296 subjects were used for the IDP deviation-PGS correlation, and 6,181 subjects were used for the Jacobian-PGS correlation. We performed a Spearman correlation analysis between the deviation scores of the IDP or Jacobian test sample and the PGS. An FDR correction was applied using the Benjamini-Hochberg method to the resulting p-values to correct for multiple comparisons. All the results of the different analyses can be found in Table 1-4.

**Supplemental Table 1.**
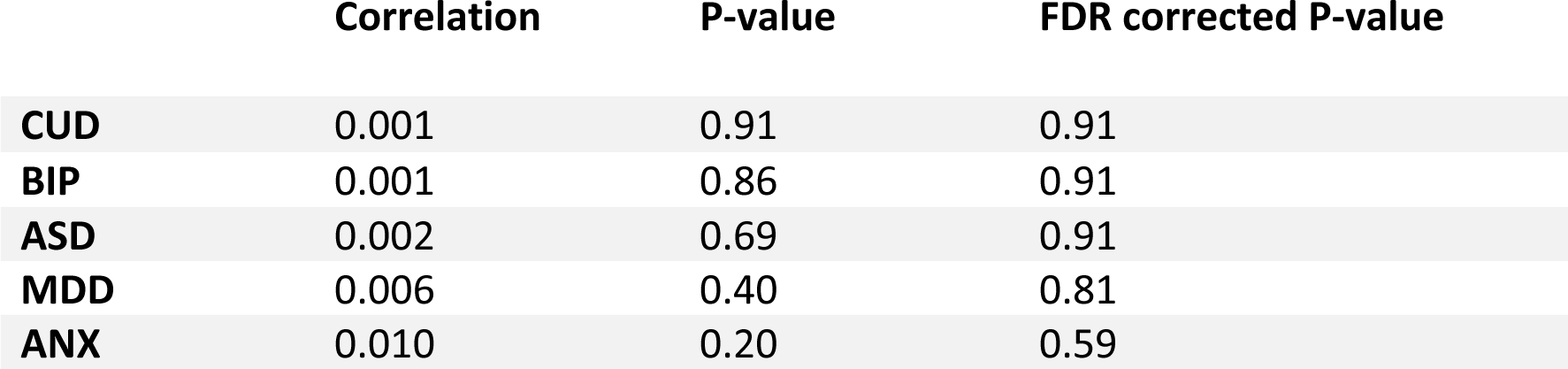
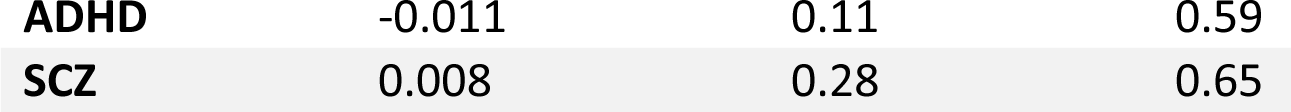
| summarizes the correlation and p-values between the different PGSs for several psychiatric disorders (CUD, BIP, ASD, MDD, ANX, ADHD, and SCZ) and the sum score for the extreme negative brain deviations (Z<-2) of the IDP normative model.

|  | Correlation | P-value | FDR corrected P-value |
| --- | --- | --- | --- |
| **CUD** | 0.001 | 0.91 | 0.91 |
| **BIP** | 0.001 | 0.86 | 0.91 |
| **ASD** | 0.002 | 0.69 | 0.91 |
| **MDD** | 0.006 | 0.40 | 0.81 |
| **ANX** | 0.010 | 0.20 | 0.59 |
| **ADHD** | -0.011 | 0.11 | 0.59 |
| **SCZ** | 0.008 | 0.28 | 0.65 |

**Supplemental Table 2.**
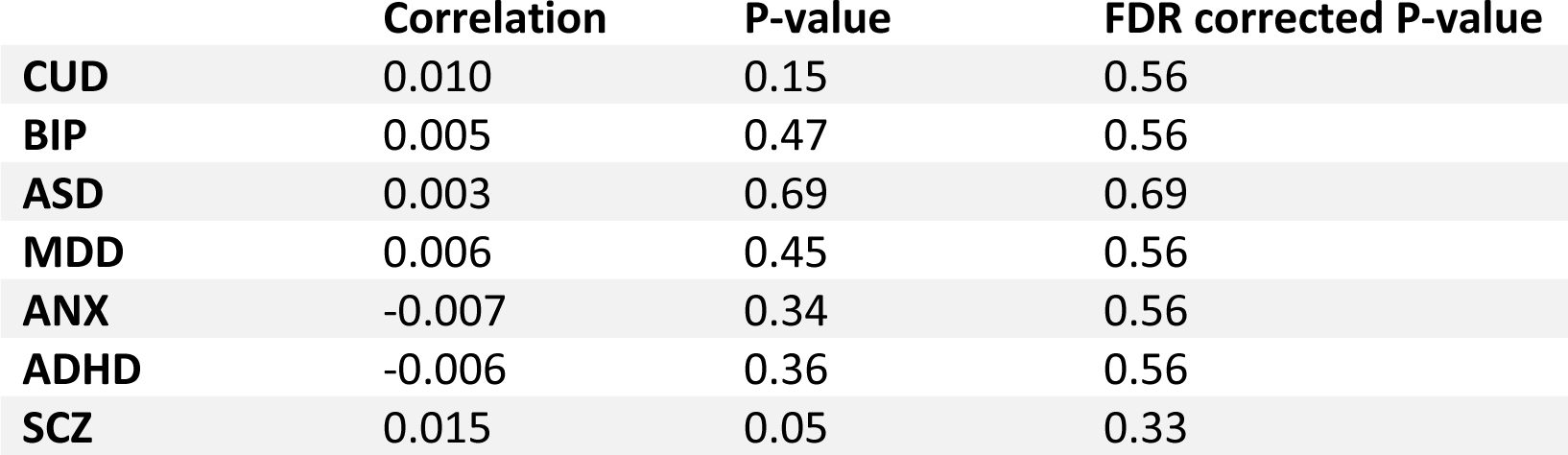
| summarizes the correlation and p-values between the different PGSs for several psychiatric disorders (CUD, BIP, ASD, MDD, ANX, ADHD, and SCZ) and the sum score for the extreme positive brain deviations (Z>2) of the IDP normative model.

|  | **Correlation** | **P-value** | **FDR corrected P-value** |
| --- | --- | --- | --- |
| **CUD** | 0.010 | 0.15 | 0.56 |
| **BIP** | 0.005 | 0.47 | 0.56 |
| **ASD** | 0.003 | 0.69 | 0.69 |
| **MDD** | 0.006 | 0.45 | 0.56 |
| **ANX** | -0.007 | 0.34 | 0.56 |
| **ADHD** | -0.006 | 0.36 | 0.56 |
| **SCZ** | 0.015 | 0.05 | 0.33 |

**Supplemental Table 3.** | summarizes the correlation and p-values between the different PGSs for several psychiatric disorders (CUD, BIP, ASD, MDD, ANX, ADHD, and SCZ) and the sum score for the extreme negative brain deviations (Z<-2) of the Jacobian normative model.

|  | **Correlation** | **P-value** | **FDR corrected P-value** |
| --- | --- | --- | --- |
| **CUD** | 0.014 | 0.27 | 0.37 |
| **BIP** | 0.019 | 0.14 | 0.25 |
| **ASD** | 0.000 | 1.00 | 1.00 |
| **MDD** | 0.020 | 0.10 | 0.23 |
| **ANX** | 0.003 | 0.77 | 0.89 |
| **ADHD** | -0.025 | 0.05 | 0.16 |
| **SCZ** | 0.025 | 0.05 | 0.16 |

**Supplemental Table 4.** | summarizes the correlation and p-values between the different PGSs for several psychiatric disorders (CUD, BIP, ASD, MDD, ANX, ADHD, and SCZ) and the sum score for the extreme positive brain deviations (Z>2) of the Jacobian normative model.

|  | **Correlation** | **P-value** | **FDR corrected P-value** |
| --- | --- | --- | --- |
| **CUD** | -0.018 | 0.13 | 0.47 |
| **BIP** | -0.009 | 0.47 | 0.67 |
| **ASD** | 0.002 | 0.90 | 0.89 |
| **MDD** | -0.020 | 0.11 | 0.47 |
| **ANX** | -0.009 | 0.48 | 0.67 |
| **ADHD** | 0.006 | 0.62 | 0.73 |
| **SCZ** | -0.015 | 0.25 | 0.58 |

## References

1. Malhotra D, Sebat J. CNVs: harbingers of a rare variant revolution in psychiatric genetics. Cell. 2012 Mar 16;148(6):1223–41.

2. Sanders SJ, Sahin M, Hostyk J, Thurm A, Jacquemont S, Avillach P, et al. A framework for the investigation of rare genetic disorders in neuropsychiatry. Nat Med. 2019 Oct;25(10):1477–87.

3. Gudmundsson OO, Walters GB, Ingason A, Johansson S, Zayats T, Athanasiu L, et al. Attention-deficit hyperactivity disorder shares copy number variant risk with schizophrenia and autism spectrum disorder. Transl Psychiatry. 2019 Oct 17;9(1):1–9.

4. Marshall CR, Howrigan DP, Merico D, Thiruvahindrapuram B, Wu W, Greer DS, et al. Contribution of copy number variants to schizophrenia from a genome-wide study of 41,321 subjects. Nat Genet. 2017 Jan;49(1):27–35.

5. Kirov G, Rees E, Walters JTR, Escott-Price V, Georgieva L, Richards AL, et al. The penetrance of copy number variations for schizophrenia and developmental delay. Biol Psychiatry. 2014 Mar 1;75(5):378–85.

6. Cooper GM, Coe BP, Girirajan S, Rosenfeld JA, Vu TH, Baker C, et al. A copy number variation morbidity map of developmental delay. Nat Genet. 2011 Sep;43(9):838–46.

7. Sanders SJ, He X, Willsey AJ, Ercan-Sencicek AG, Samocha KE, Cicek AE, et al. Insights into Autism Spectrum Disorder Genomic Architecture and Biology from 71 Risk Loci. Neuron. 2015 Sep 23;87(6):1215–33.

8. Fromer M, Pocklington AJ, Kavanagh DH, Williams HJ, Dwyer S, Gormley P, et al. De novo mutations in schizophrenia implicate synaptic networks. Nature. 2014 Feb 13;506(7487):179–84.

9. International Schizophrenia Consortium. Rare chromosomal deletions and duplications increase risk of schizophrenia. Nature. 2008 Sep 11;455(7210):237–41.

10. Walsh T, McClellan JM, McCarthy SE, Addington AM, Pierce SB, Cooper GM, et al. Rare structural variants disrupt multiple genes in neurodevelopmental pathways in schizophrenia. Science. 2008 Apr 25;320(5875):539–43.

11. Bearden CE, Forsyth JK. The many roads to psychosis: recent advances in understanding risk and mechanisms. F1000Res. 2018 Dec 3;7:F1000 Faculty Rev-1883.

12. Singh T, Poterba T, Curtis D, Akil H, Al Eissa M, Barchas JD, et al. Rare coding variants in ten genes confer substantial risk for schizophrenia. Nature. 2022 Apr;604(7906):509–16.

13. Lionel AC, Crosbie J, Barbosa N, Goodale T, Thiruvahindrapuram B, Rickaby J, et al. Rare copy number variation discovery and cross-disorder comparisons identify risk genes for ADHD. Sci Transl Med. 2011 Aug 10;3(95):95ra75.

14. Moreau CA, Urchs SGW, Kuldeep K, Orban P, Schramm C, Dumas G, et al. Mutations associated with neuropsychiatric conditions delineate functional brain connectivity dimensions contributing to autism and schizophrenia. Nat Commun. 2020 Oct 19;11(1):5272.

15. Douard E, Zeribi A, Schramm C, Tamer P, Loum MA, Nowak S, et al. Effect Sizes of Deletions and Duplications on Autism Risk Across the Genome. Am J Psychiatry. 2021 Jan 1;178(1):87–98.

16. Stefansson H, Meyer-Lindenberg A, Steinberg S, Magnusdottir B, Morgen K, Arnarsdottir S, et al. CNVs conferring risk of autism or schizophrenia affect cognition in controls. Nature. 2014 Jan;505(7483):361–6.

17. Lee PH, Anttila V, Won H, Feng YCA, Rosenthal J, Zhu Z, et al. Genomic Relationships, Novel Loci, and Pleiotropic Mechanisms across Eight Psychiatric Disorders. Cell. 2019 Dec 12;179(7):1469–1482.e11.

18. Andrews T, Meader S, Vulto-van Silfhout A, Taylor A, Steinberg J, Hehir-Kwa J, et al. Gene networks underlying convergent and pleiotropic phenotypes in a large and systematically-phenotyped cohort with heterogeneous developmental disorders. PLoS Genet. 2015 Mar;11(3):e1005012.

19. Moreau C, Huguet G, Urchs S, Douard E, Sharmarke H, Orban P, et al. The general impact of haploinsufficiency on brain connectivity underlies the pleiotropic effect of neuropsychiatric CNVs [Internet]. medRxiv; 2020 [cited 2023 Aug 14]. p. 2020.03.18.20038505. Available from: https://www.medrxiv.org/content/10.1101/2020.03.18.20038505v1

20. Richetto J, Meyer U. Epigenetic Modifications in Schizophrenia and Related Disorders: Molecular Scars of Environmental Exposures and Source of Phenotypic Variability. Biological Psychiatry. 2021 Feb 1;89(3):215–26.

21. Boen R, Kaufmann T, van der Meer D, Frei O, Agartz I, Ames D, et al. Beyond the Global Brain Differences: Intra-individual Variability Differences in 1q21.1 Distal and 15q11.2 BP1-BP2 Deletion Carriers. Biol Psychiatry. 2023 Sep 1;S0006–3223(23)01530-5.

22. Marquand AF, Rezek I, Buitelaar J, Beckmann CF. Understanding Heterogeneity in Clinical Cohorts Using Normative Models: Beyond Case-Control Studies. Biol Psychiatry. 2016 Oct 1;80(7):552–61.

23. Marquand AF, Kia SM, Zabihi M, Wolfers T, Buitelaar JK, Beckmann CF. Conceptualizing mental disorders as deviations from normative functioning. Mol Psychiatry. 2019;24(10):1415–24.

24. Rutherford S, Fraza C, Dinga R, Kia SM, Wolfers T, Zabihi M, et al. Charting brain growth and aging at high spatial precision. Baker CI, Taschler B, Esteban O, Constable T, editors. eLife. 2022 Feb 1;11:e72904.

25. Rutherford S, Barkema P, Tso IF, Sripada C, Beckmann CF, Ruhe HG, et al. Evidence for Embracing Normative Modeling [Internet]. Neuroscience; 2022 Nov [cited 2023 Feb 6]. Available from: http://biorxiv.org/lookup/doi/10.1101/2022.11.14.516460

26. Bučková BR, Fraza C, Rehák R, Kolenič M, Beckmann C, Španiel F, et al. Using normative models pre-trained on cross-sectional data to evaluate longitudinal changes in neuroimaging data [Internet]. bioRxiv; 2023 [cited 2023 Aug 14]. p. 2023.06.09.544217. Available from: https://www.biorxiv.org/content/10.1101/2023.06.09.544217v1

27. Bethlehem R a. I, Seidlitz J, White SR, Vogel JW, Anderson KM, Adamson C, et al. Brain charts for the human lifespan. Nature. 2022 Apr;604(7906):525–33.

28. Wolfers T, Doan NT, Kaufmann T, Alnæs D, Moberget T, Agartz I, et al. Mapping the Heterogeneous Phenotype of Schizophrenia and Bipolar Disorder Using Normative Models. JAMA Psychiatry. 2018 Nov 1;75(11):1146–55.

29. Wolfers T, Beckmann CF, Hoogman M, Buitelaar JK, Franke B, Marquand AF. Individual differences v. the average patient: mapping the heterogeneity in ADHD using normative models. Psychol Med. 2020 Jan;50(2):314–23.

30. Zabihi M, Floris DL, Kia SM, Wolfers T, Tillmann J, Arenas AL, et al. Fractionating autism based on neuroanatomical normative modeling. Transl Psychiatry. 2020 Nov 6;10(1):1– 10.

31. Pinaya WHL, Scarpazza C, Garcia-Dias R, Vieira S, Baecker L, F da Costa P, et al. Using normative modelling to detect disease progression in mild cognitive impairment and Alzheimer’s disease in a cross-sectional multi-cohort study. Sci Rep. 2021 Aug 3;11(1):15746.

32. Owen MJ, O’Donovan MC. Schizophrenia and the neurodevelopmental continuum:evidence from genomics. World Psychiatry. 2017 Oct;16(3):227–35.

33. Fraza C, Zabihi M, Beckmann CF, Marquand AF. The Extremes of Normative Modelling [Internet]. bioRxiv; 2022 [cited 2023 Jan 23]. p. 2022.08.23.505049. Available from: https://www.biorxiv.org/content/10.1101/2022.08.23.505049v1

34. Taylor JR, Williams N, Cusack R, Auer T, Shafto MA, Dixon M, et al. The Cambridge Centre for Ageing and Neuroscience (Cam-CAN) data repository: Structural and functional MRI, MEG, and cognitive data from a cross-sectional adult lifespan sample. NeuroImage. 2017 Jan 1;144:262–9.

35. Van Essen DC, Smith SM, Barch DM, Behrens TEJ, Yacoub E, Ugurbil K. The WU-Minn Human Connectome Project: An overview. NeuroImage. 2013 Oct 15;80:62–79.

36. Marcus DS, Fotenos AF, Csernansky JG, Morris JC, Buckner RL. Open Access Series of Imaging Studies (OASIS): Longitudinal MRI Data in Nondemented and Demented Older Adults. J Cogn Neurosci. 2010 Dec;22(12):2677–84.

37. Satterthwaite TD, Elliott MA, Ruparel K, Loughead J, Prabhakaran K, Calkins ME, et al. Neuroimaging of the Philadelphia Neurodevelopmental Cohort. Neuroimage. 2014 Feb 1;86:544–53.

38. Alfaro-Almagro F, Jenkinson M, Bangerter NK, Andersson JLR, Griffanti L, Douaud G, et al. Image processing and Quality Control for the first 10,000 brain imaging datasets from UK Biobank. NeuroImage. 2018 Feb 1;166:400–24.

39. McCarthy P. Funpack. 2020.

40. Monté-Rubio GC, Falcón C, Pomarol-Clotet E, Ashburner J. A comparison of various MRI feature types for characterizing whole brain anatomical differences using linear pattern recognition methods. NeuroImage. 2018 Sep 1;178:753–68.

41. Sønderby IE, van der Meer D, Moreau C, Kaufmann T, Walters GB, Ellegaard M, et al. 1q21.1 distal copy number variants are associated with cerebral and cognitive alterations in humans. Transl Psychiatry. 2021 Mar 22;11(1):1–16.

42. Modenato C, Martin-Brevet S, Moreau CA, Rodriguez-Herreros B, Kumar K, Draganski B, et al. Lessons Learned From Neuroimaging Studies of Copy Number Variants: A Systematic Review. Biological Psychiatry. 2021 Nov 1;90(9):596–610.

43. Fraza CJ, Dinga R, Beckmann CF, Marquand AF. Warped Bayesian linear regression for normative modelling of big data. Neuroimage. 2021 Dec 15;245:118715.

44. Nygaard V, Rødland EA, Hovig E. Methods that remove batch effects while retaining group differences may lead to exaggerated confidence in downstream analyses. Biostatistics. 2016 Jan 1;17(1):29–39.

45. Bayer JMM, Thompson PM, Ching CRK, Liu M, Chen A, Panzenhagen AC, et al. Site effects how-to and when: An overview of retrospective techniques to accommodate site effects in multi-site neuroimaging analyses. Frontiers in Neurology [Internet]. 2022 [cited 2024 Feb 5];13. Available from: https://www.frontiersin.org/journals/neurology/articles/10.3389/fneur.2022.923988

46. Bayer JMM, Dinga R, Kia SM, Kottaram AR, Wolfers T, Lv J, et al. Accommodating site variation in neuroimaging data using normative and hierarchical Bayesian models. NeuroImage. 2022 Dec 1;264:119699.

47. Kendall KM, Rees E, Escott-Price V, Einon M, Thomas R, Hewitt J, et al. Cognitive Performance Among Carriers of Pathogenic Copy Number Variants: Analysis of 152,000 UK Biobank Subjects. Biological Psychiatry. 2017 Jul 15;82(2):103–10.

48. Kendall KM, Bracher-Smith M, Fitzpatrick H, Lynham A, Rees E, Escott-Price V, et al. Cognitive performance and functional outcomes of carriers of pathogenic copy number variants: analysis of the UK Biobank. The British Journal of Psychiatry. 2019 May;214(5):297–304.

49. Crawford K, Bracher-Smith M, Owen D, Kendall KM, Rees E, Pardiñas AF, et al. Medical consequences of pathogenic CNVs in adults: analysis of the UK Biobank. J Med Genet. 2019 Mar;56(3):131–8.

50. Clinical phenotype of the recurrent 1q21.1 copy-number variant - ScienceDirect [Internet]. [cited 2023 Aug 28]. Available from: https://www.sciencedirect.com/science/article/pii/S1098360021043306?via%3Dihub

51. Stefansson H, Rujescu D, Cichon S, Pietiläinen OPH, Ingason A, Steinberg S, et al. Large recurrent microdeletions associated with schizophrenia. Nature. 2008 Sep;455(7210):232–6.

52. Green EK, Rees E, Walters JTR, Smith KG, Forty L, Grozeva D, et al. Copy number variation in bipolar disorder. Mol Psychiatry. 2016 Jan;21(1):89–93.

53. Sønderby IE, Gústafsson Ó, Doan NT, Hibar DP, Martin-Brevet S, Abdellaoui A, et al. Dose response of the 16p11.2 distal copy number variant on intracranial volume and basal ganglia. Mol Psychiatry. 2020;25(3):584–602.

54. Modenato C, Kumar K, Moreau C, Martin-Brevet S, Huguet G, Schramm C, et al. Effects of eight neuropsychiatric copy number variants on human brain structure. Transl Psychiatry. 2021 Jul 20;11(1):399.

55. Sripada C, Angstadt M, Rutherford S, Taxali A, Shedden K. Toward a “treadmill test” for cognition: Improved prediction of general cognitive ability from the task activated brain. Human Brain Mapping. 2020 May 1;41.

56. Bernier R, Steinman KJ, Reilly B, Wallace AS, Sherr EH, Pojman N, et al. Clinical phenotype of the recurrent 1q21.1 copy-number variant. Genet Med. 2016 Apr;18(4):341–9.

57. Brunetti-Pierri N, Berg JS, Scaglia F, Belmont J, Bacino CA, Sahoo T, et al. Recurrent reciprocal 1q21.1 deletions and duplications associated with microcephaly or macrocephaly and developmental and behavioral abnormalities. Nat Genet. 2008 Dec;40(12):1466–71.

58. Ge T, Chen CY, Ni Y, Feng YCA, Smoller JW. Polygenic prediction via Bayesian regression and continuous shrinkage priors. Nat Commun. 2019 Apr 16;10(1):1776.

59. Genetic liability to major psychiatric disorders contributes to multi-faceted quality of life outcomes in children and adults | medRxiv [Internet]. [cited 2024 May 31]. Available from: https://www.medrxiv.org/content/10.1101/2023.01.17.23284645v1.full

60. Li J, Bzdok D, Chen J, Tam A, Ooi LQR, Holmes AJ, et al. Cross-ethnicity/race generalization failure of behavioral prediction from resting-state functional connectivity. Science Advances. 2022 Mar 16;8(11):eabj1812.

61. LaMontagne PJ, Benzinger TL, Morris JC, Keefe S, Hornbeck R, Xiong C, et al. OASIS-3: Longitudinal Neuroimaging, Clinical, and Cognitive Dataset for Normal Aging and Alzheimer Disease [Internet]. medRxiv; 2019 [cited 2024 May 22]. p. 2019.12.13.19014902. Available from: https://www.medrxiv.org/content/10.1101/2019.12.13.19014902v1

62. Writing Committee for the ENIGMA-CNV Working Group, van der Meer D, Sønderby IE, Kaufmann T, Walters GB, Abdellaoui A, et al. Association of Copy Number Variation of the 15q11.2 BP1-BP2 Region With Cortical and Subcortical Morphology and Cognition. JAMA Psychiatry. 2020 Apr 1;77(4):420–30.

63. Huguet G, Schramm C, Douard E, Jiang L, Labbe A, Tihy F, et al. Measuring and Estimating the Effect Sizes of Copy Number Variants on General Intelligence in Community-Based Samples. JAMA Psychiatry. 2018 May 1;75(5):447–57.

64. Research Domain Criteria (RDoC): Toward a New Classification Framework for Research on Mental Disorders | American Journal of Psychiatry [Internet]. [cited 2023 Aug 31]. Available from: https://ajp.psychiatryonline.org/doi/10.1176/appi.ajp.2010.09091379?url_ver=Z39.88-2003&rfr_id=ori:rid:crossref.org&rfr_dat=cr_pub%20%200pubmed

65. Szecówka K, Misiak B, Łaczmańska I, Frydecka D, Moustafa AA. Copy Number Variations and Schizophrenia. Mol Neurobiol. 2023 Apr 1;60(4):1854–64.

66. Xu J, Liu N, Polemiti E, Garcia-Mondragon L, Tang J, Liu X, et al. Effects of urban living environments on mental health in adults. Nat Med. 2023 Jun 15;1–12.

67. Di Biase MA, Tian YE, Bethlehem RAI, Seidlitz J, Alexander-Bloch AaronF, Yeo BTT, et al. Mapping human brain charts cross-sectionally and longitudinally. Proceedings of the National Academy of Sciences. 2023 May 16;120(20):e2216798120.

68. Henrich J, Heine SJ, Norenzayan A. The weirdest people in the world? Behav Brain Sci. 2010 Jun;33(2–3):61–83; discussion 83-135.

69. Holz NE, Zabihi M, Kia SM, Monninger M, Aggensteiner PM, Siehl S, et al. A stable and replicable neural signature of lifespan adversity in the adult brain. Nat Neurosci. 2023 Aug 21;1–10.

70. Kia SM, Huijsdens H, Rutherford S, Dinga R, Wolfers T, Mennes M, et al. Federated Multi-Site Normative Modeling using Hierarchical Bayesian Regression. bioRxiv. 2021 May 30;2021.05.28.446120.

71. Fawns-Ritchie C, Deary IJ. Reliability and validity of the UK Biobank cognitive tests. PLOS ONE. 2020 Apr 20;15(4):e0231627.

72. Lyall DM, Cullen B, Allerhand M, Smith DJ, Mackay D, Evans J, et al. Cognitive Test Scores in UK Biobank: Data Reduction in 480,416 Participants and Longitudinal Stability in 20,346 Participants. PLOS ONE. 2016 Apr 25;11(4):e0154222.

73. Demontis D, Walters GB, Athanasiadis G, Walters R, Therrien K, Nielsen TT, et al. Genome-wide analyses of ADHD identify 27 risk loci, refine the genetic architecture and implicate several cognitive domains. Nat Genet. 2023 Feb;55(2):198–208.

74. Matoba N, Liang D, Sun H, Aygün N, McAfee JC, Davis JE, et al. Common genetic risk variants identified in the SPARK cohort support DDHD2 as a candidate risk gene for autism. Transl Psychiatry. 2020 Aug 3;10(1):265.

75. Wray NR, Ripke S, Mattheisen M, Trzaskowski M, Byrne EM, Abdellaoui A, et al. Genome-wide association analyses identify 44 risk variants and refine the genetic architecture of major depression. Nat Genet. 2018 May;50(5):668–81.

76. Otowa T, Hek K, Lee M, Byrne EM, Mirza SS, Nivard MG, et al. Meta-analysis of genome-wide association studies of anxiety disorders. Mol Psychiatry. 2016 Oct;21(10):1391–9.

77. Purves KL, Coleman JRI, Meier SM, Rayner C, Davis KAS, Cheesman R, et al. A major role for common genetic variation in anxiety disorders. Mol Psychiatry. 2020 Dec;25(12):3292–303.

78. Trubetskoy V, Pardiñas AF, Qi T, Panagiotaropoulou G, Awasthi S, Bigdeli TB, et al. Mapping genomic loci implicates genes and synaptic biology in schizophrenia. Nature. 2022 Apr;604(7906):502–8.

79. Mullins N, Forstner AJ, O’Connell KS, Coombes B, Coleman JRI, Qiao Z, et al. Genome-wide association study of more than 40,000 bipolar disorder cases provides new insights into the underlying biology. Nat Genet. 2021 Jun;53(6):817–29.

80. Johnson EC, Demontis D, Thorgeirsson TE, Walters RK, Polimanti R, Hatoum AS, et al. A large-scale genome-wide association study meta-analysis of cannabis use disorder. Lancet Psychiatry. 2020 Dec;7(12):1032–45.

